# Biomedical Large Language Models and Prompt Engineering for Causality Assessment of Individual Case Safety Reports in Pharmacovigilance

**DOI:** 10.64898/2026.02.19.26346467

**Authors:** Nicole Sonne Heckmann, Despoina Georgia Papoutsi, Maria Antonietta Barbieri, Vera Battini, Søren Norlin Mølgaard, Simon Ørum Schmidt, Lars Melskens, Maurizio Sessa

## Abstract

**Background:** Biomedical Large Language Models (LLMs) combined with prompt engineering offer domain-specific reasoning, yet their application to individual-level causality assessment remains unexplored. This study evaluated five combinations of biomedical LLMs, prompting strategies, and causality algorithms by comparing their agreement with two human expert evaluators.

**Research design and methods:** A total of 150 Individual Case Safety Reports (ICSRs) were analyzed: 140 reports from Food and Drug Administration Adverse Event Reporting System (FAERS), and 10 myocarditis/pericarditis ICSRs from Vaccine AERS (VAERS). Assessments were conducted using the Naranjo and WHO-UMC algorithms. Biomedical LLMs tested included TinyLlama 1.1B, Medicine LLaMA-3 8B, and MedLLaMA v20, combined with Chain-of-Thought (CoT) or Decomposition prompting. Agreement was measured using Gwet’s Agreement Coefficient 1 (AC1) and percentage agreement, alongside performance metrics and qualitative error analysis.

**Results:** The Medicine LLaMA-3 8B-Naranjo-CoT combination achieved the highest agreement with human assessors for the final classification of causality (64%). Biomedical LLMs demonstrated low inter-rater agreement on critical items of causality assessment such as identification of listed AE, temporal plausibility, alternative causes, and objective evidence of AEs. Frequent model failures included irrelevant responses.

**Conclusions:** Biomedical LLMs showed improved performance over general purpose models previously tested but remain suboptimal for reliable causality assessment of ICSRs.

## 1. Introduction

One of the key processes in pharmacovigilance is causality assessment of individual case safety reports (ICSRs), which aims to determine the likelihood of a causal association between drugs or vaccines and adverse events/adverse events following immunization (AEs/AEFIs) in a specific case [1]. Reliable and timely safety assessments in pharmacovigilance require robust case-by-case causality evaluation methods, even when regulatory authorities face substantial surges in reporting [2–5]. Manual causality assessment has become increasingly unsustainable, revealing significant limitations in current workflows and underscoring the need for innovative solutions to manage large volumes of ICSRs [6]. Existing approaches, that typically combine expert judgment, rule-based algorithms, and statistical techniques, are inherently time- and resource-intensive [7,8]. Large Language Models (LLMs) have demonstrated impressive capabilities in interpreting and reasoning over clinical text across several pharmacovigilance applications [6]. However, performance is still suboptimal. A recent study by Abate et al. highlighted general-purpose LLMś limitations in individual-level causality assessment, suggesting that suboptimal performance may stem from inadequate prompt strategies and the lack of domain-specific training data [8]. Currently, there is a notable lack of studies investigating the potential of biomedical literature-trained LLMs combined with state-of-the-art prompt engineering techniques for individual-level causality assessment. Therefore, this study aimed to: (1) evaluate the feasibility of using biomedical LLMs with various prompt engineering strategies for individual-level causality assessment by calculating inter-rater and reasoning agreement between the biomedical LLMs and human experts; (2) analyze the errors and inconsistencies of biomedical LLMs during individual-level causality assessment.

## 2. Methods

### 2.1 Data Sources

A total of 140 cases were extracted from the U.S. Food and Drug Administration Adverse Event Reporting System (FAERS). Data was accessed via the OpenFDA portal as quarterly ASCII files (Quarter 4 2023 and Quarter 1 2024). Record-linked case identifiers were used to integrate information from the seven datasets in the quarterly files, including patient demographics, drug exposure, reported AEs, indications, outcomes, and reporter details. ICSRs were randomly sampled if they had reported as suspected drugs across six predefined therapeutic categories: (1) newly approved drugs (i.e., semaglutide, teprotumumab, tirzepatide); (2) gene and cell therapies (Anatomical Therapeutic Chemical classification (ATC) code L01XL); (3) commonly prescribed drugs in the United States for highly prevalent conditions (i.e., pain, mental health, and acid-related disorders) [9]; (4) drugs requiring special safety monitoring (i.e., ciprofloxacin, lamotrigine, isoniazid); (5) orphan drugs for rare diseases (i.e., agalsidase beta, nusinersen); and (6) controlled substances (i.e., oxycodone, alprazolam). Sampling was performed using the R function *sample* (R version 3.6.2). Additionally, 10 clinically confirmed cases of myocarditis and pericarditis following vaccination with Comirnaty® (Pfizer-BioNTech), Spikevax® (Moderna), or Janssen® (Johnson & Johnson), reported to the Vaccine Adverse Event Reporting System (VAERS) up to 28^th^ July 2021, were included. The 10 VAERS cases were sampled among those previously analyzed in 2023 by Abate et al., [8] using the R function *sample* (R version 3.6.2). For these cases, clinical evaluation was already conducted by a multidisciplinary panel comprising cardiologists, epidemiologists, and virologists.

### 2.2 Data Preparation

All FAERS tables were record-linked according to the relational structure described by Khaleel et al. [10], enabling reconstruction of complete, case-level records across demographic, drug, reaction, outcome, and reporting tables. For each case, regulatory identifiers, patient characteristics, product information, exposure details, AEs, outcomes, and reporting metadata were collected (Supplementary Table 1). All cases were then manually reviewed to identify potential duplicate reports, by considering overlapping identifiers, product attributes, event dates, and clinical details as described by Kibura et al., [11].

### 2.3 Causality Assessment Algorithm Prioritization

The Naranjo and World Health Organization-Uppsala Monitoring Centre (WHO-UMC) algorithms were used for causality assessment as they are two of the most used algorithms worldwide [12].

#### 2.3.1 Naranjo algorithm

The Naranjo Adverse Drug Reaction Probability Scale is one of the most widely used tools for assessing the likelihood that an AE is drug related [12]. It consists of ten structured questions with weighted responses (–1, 0, +1, +2), and the total score classifies causality as doubtful (≤0), possible (1-4), probable (5-8), or definite (≥9). The ten questions evaluate key drug-event relationships, including prior evidence, temporal association, dechallenge and rechallenge outcomes, alternative causes, dose-response relationships, prior patient history, and objective clinical or laboratory confirmation. Details about the Naranjo algorithm are provided elsewhere [13].

#### 2.3.2 WHO-UMC algorithm

The WHO-UMC algorithm is a structured approach for individual-level causality assessment in AEFI, following a four-step process: eligibility, checklist, algorithm, and classification. Eligibility confirms that drug or vaccine exposure preceded the AEFI and is supported by clinical and diagnostic information. The checklist evaluates alternative explanations, existing evidence for or against a causal association, and contributory factors such as comorbidities or concomitant medications. Events with a plausible causal link are then assessed using the algorithm and classified as consistent with causal association, indeterminate, inconsistent, or unclassifiable. Details about the WHO-UMC algorithm are provided elsewhere [14].

### 2.4 Biomedical Large Language Model Prioritization

TinyLlama 1.1B (Developer: Afrideva), Medicine LLaMA-3 8B (Developer: QuantFactory), and MedLLaMA v20 (Developer: JL42) were selected as the biomedical LLMs evaluated in this study. Model selection was based on overall performance as reported by the Open LLM Leaderboard hosted on Hugging Face (https://huggingface.co/spaces/open-llm-leaderboard/open_llm_leaderboard#/). For the performance assessment, standardized evaluation metrics where considered including IFEval (Instruction-Following Evaluation), which measures adherence to explicit user instructions; BBH (Big-Bench Hard), which assesses advanced reasoning across challenging tasks; MATH, which evaluates mathematical and symbolic reasoning accuracy; and GPQA (Graduate-level Google-Proof Q&A), which probes expert-level reasoning on difficult, domain-agnostic questions [15]. Available biomedical LLMs at the time of evaluations were ranked using the aggregate average score across these benchmarks, and the top three performing biomedical LLMs as of January 2025 were selected for subsequent analysis. For the selected biomedical LLMs, inference was conducted using off-the-shelf hyperparameter configurations. The generation settings included a temperature of 0.8, top-K sampling of 40, a repetition penalty of 1.0, top-P (nucleus) sampling of 0.95, min-P sampling of 0.05, and a fixed random seed to ensure reproducibility (i.e., 1234). The maximum context length differed across biomedical LLMs, with TinyLlama 1.1B (Developer: Afrideva) supporting up to 2,048 tokens, and both Medicine LLaMA-3 8B (Developer: QuantFactory) and MedLLaMA v20 (Developer: JL42) supporting up to 4,096 tokens and had 8B parameters. The three biomedical LLMs were open source at the time we conducted the study, and the cut-off dates were December 2023, June 2024 and May 2024 for TinyLlama 1.1B[16], LLaMA-3 8B[17], and MedLLaMA v20 [18], respectively.

### 2.5 Prompt Strategy Prioritization

We consulted the public performance benchmark and leaderboard reported by the LLM-Eval project (https://llm-eval.github.io/pages/leaderboard/pe.html), which compares prompt strategies based on standardized evaluation outcomes as described by Zhu et al., [19]. Based on evidence from this benchmark, Chain-of-Thought (CoT) prompting was selected as the most promising strategy. Decomposition was implemented as the second prioritized prompt strategy based on evidence from a review on prompt engineering [20]. All prompts adhered to the Concise, Logical, Explicit, Adaptive, and Reflective (CLEAR) principles [21].

### 2.6 Prompts and Causality Assessment

The prompt templates used in this study are provided in Table 1. The prompts were developed by the lead author in collaboration with a natural language processing (NLP) engineer. Causality assessment of ICSRs retrieved from FAERS was performed by expert assessors, who served as the gold-standard reference for evaluation. Specifically, the two assessors were a pharmacist with expertise in pharmacovigilance and a senior medical doctor at Novo Nordisk, who evaluated 140 cases between February and May 2025 using both the Naranjo and WHO-UMC algorithms. For each algorithm, individual question-level responses were collected with the supporting clinical information and rationale provided by the assessors and systematically recorded in an Excel file (Supplementary Table 2). The outputs generated by each biomedical LLM-prompt-algorithm combination, including individual question responses and the corresponding supporting clinical information or rationale produced by the models, were collected and stored (Supplementary Table 2).

**Table 1.** Prompt Templates.

| *Prompt Text* |  |  |
| --- | --- | --- |
|  | *Chain-of-Thoughts* | *Decomposition* |
| **General instructions** | Above I have provided the details of a pharmacovigilance case involving the adverse event of X after the administration of the drug Y. The primary suspect is X and the adverse event that I would like to assess causality is Y.<br>I would like you to perform a step-by-step causality assessment for the reported AEs using the Naranjo algorithm. Apply logical reasoning to evaluate each question in sequence. Consider all available data and explain how each answer contributes to the overall causality assessment.<br>Based on the responses that you will give, calculate the final causality score separately for the AEs. Summarize each score and interpret it using the following categories: Definite ($\geq 9$), Probable (5-8), Possible (1-4), Doubtful (0). | Above I have provided the details of a pharmacovigilance case involving the adverse event of X after the administration of the drug Y. The primary suspect is X and the adverse event that I would like to assess causality is Y. For each question in the Naranjo Algorithm, respond based on the case information provided. Each answer should include specific reasoning related to the reported adverse event. Once all questions have been answered individually, combine the responses to calculate a causality score for each AE.<br>Based on the responses that you will give, calculate the final causality score separately for the AEs. Summarize each score and interpret it using the following categories: Definite ($\geq 9$), Probable (5-8), Possible (1-4), Doubtful (0). If the score is close to a category boundary, discuss any uncertainties or conflicting evidence that might affect the classification. |
| **Question 1** | Are there previous conclusive reports on the specific AE after the administration of the suspected drug?<br>Reasoning: Based on available medical literature, pharmacovigilance reports, the product package insert or at the summary of product characteristics (SmPC). Analyze whether the suspected drug has been previously associated with this specific AE. If conclusive evidence exists, explain how it supports causality. | Are there previous conclusive reports on the specific AE after the administration of the suspected drug?<br>Answer this question primarily based on the FAERS case data provided. If additional evidence from literature or pharmacovigilance reports is relevant, discuss how it supports or contradicts the FAERS findings. Explain if there is evidence supporting a link between suspected drug and the AE. |
| **Question 2** | Did AE appear after the suspected drug was administered?<br>Reasoning: given the reported timeline and the known route of administration, consider the likelihood that the reaction was temporally related to suspected drug administration. Analyze if the timing is consistent with expected AE and answer whether the AE appeared immediately, shortly after, or with a delayed onset. | Did AE appear after suspected drug administration in this case? Analyze the temporal relationship between drug administration and the onset of AE. If timing information is clear, explain whether the AE occurred immediately, shortly after, or with a delayed onset, and how this affects causality. |
| **Question 3** | Did AE improve when the suspected drug was discontinued, or a specific antagonist was | Did AE improve after discontinuing suspected drug or with the administration of a specific |
|  | administered?<br>Reasoning: Search for dechallenge data in the case prompt. Discuss whether improvement upon discontinuation would have provided stronger causality evidence. If no rechallenge data is available, discuss why rechallenge is important and how the absence of this information affects the causality score. | antagonist? Search for dechallenge data. If certain information is unavailable, discuss how this uncertainty impacts the causality assessment. |
| **Question 4** | Did AE reappear when the suspected drug was readministered?<br>Reasoning: search for rechallenge data in the case provided. If marked as “not available,” reason through how the absence of this information affects the analysis. Would rechallenge data have strengthened the causality score? | Did AE reappear upon re-administration of the suspected drug? Search for rechallenge data. If certain information is unavailable, discuss how this uncertainty impacts the causality assessment. |
| **Question 5** | Are there alternative causes (other than suspected drug) that could on their own cause AE?<br>Reasoning: Examine any possible alternative explanations, such as other medications, underlying health conditions, or external factors, that could independently cause these symptoms. If none are identified, note this as supporting causality. If alternative causes are identified, explain how they affect the likelihood that the suspected drug caused the event. | Are there any alternative causes (other than suspected drug) that could independently explain AE? Consider whether other medications, underlying conditions, or environmental factors could independently explain the AE. |
| **Question 6** | Did AE appear when a placebo was given?<br>Reasoning: Search to see if placebo data is available in the case provided. If not found, reflect on how the absence of a placebo test impacts causality determination. Consider the limitations of assessing causality without this control measure. | Did AE reappear when a placebo was administered? Search for placebo data. If no information is available, explain the implications of this missing data on the causality assessment. |
| **Question 7** | Was the suspected drug detected in any body fluid in toxic concentrations? This question applies specifically to dose dependent adverse reactions when blood, urine, tissue or other specimen concentrations of the medicine are available.<br>Reasoning: search for toxicology data in the case provided. If not found, discuss how the lack of such data affects the causality score. If this information had been available, it might have provided more clarity on the association with the suspected drug. | Was the suspected drug detected in any body fluid at toxic concentrations? Search for toxicology data. If certain information is unavailable, discuss how this uncertainty impacts the causality assessment. |
| **Question 8** | Were AE more severe when the dose was | Was AE more severe when the suspected drug |
|  | increased or less severe when the dose was decreased?<br>Reasoning: search in the case provided to see if the drug has been administered in different doses. Given that the suspected drug was administered in two different doses, analyze whether there is any indication that symptoms varied with dose changes, even if data on dose-specific reactions is limited. | dose was increased or less severe when the dose was decreased? Analyze the possible dose-response relationship in this case. Assess whether the severity of the AE increased with higher doses or decreased when the dose was reduced. If a dose-response relationship is present, explain how it supports causality. |
| **Question 9** | Did the patient have a similar AE to the same or similar drugs in any previous exposure?<br>Reasoning: Consider any history of similar reactions to suspected drug or related medications, if available, and discuss how this would influence causality if relevant prior reactions existed. Consider whether the patient had prior adverse reactions to the same drug in previous exposures or to a drug in the same pharmacological class. If a positive history exists, this strengthens causality. If no history is available, discuss how this affects the overall assessment. | Did the patient experience similar AE with the suspected drug or similar drugs in previous exposures? Discuss the relevance of such a history if available. If a positive history is found, explain how it strengthens the causality assessment. |
| **Question 10** | Was the AE confirmed by any objective evidence? Reasoning: identify whether there is any medical imaging, laboratory tests, or clinical documentation confirming the reported adverse event. If objective evidence is available, discuss how it reinforces causality. Discuss how objective confirmation would impact the assessment. | Is there any objective evidence confirming the occurrence of the AE? Look for medical tests, lab results, imaging, or clinical observations that validate the reported adverse event. If objective evidence is available, explain how it strengthens the causality assessment. |

### 2.7 Study Outcomes

The primary outcome was inter-rater agreement and adherence between biomedical LLM-generated causality assessments and human expert assessments across combinations of prompt engineering strategies and established causality assessment frameworks. Inter-rater agreement was evaluated at two levels: (i) the scores assigned to individual algorithm questions and (ii) the underlying reasoning provided for each question within the causality assessment frameworks. Adherence, defined as agreement in the final causality classification, was derived from the aggregation of individual question-level responses within each causality assessment. The secondary outcome was the identification of errors and inconsistencies in biomedical LLM-generated outputs. Inconsistencies were defined as contradictory or unstable outputs across similar prompts or within a single response. Errors were classified into predefined categories, including deviations from algorithmic instructions, prompt echoing [22], lack of justification or incomplete answers [23], instructional hallucinations or inconsistencies [24], and self-dialogue loops [25].

### 2.8 Data Analysis & Software

A descriptive analysis was conducted to summarize sex, age, and the number of AEs among the ICSRs. Confusion matrices were generated to visualize agreement between biomedical LLM-generated assessments and human expert assessments. Key performance metrics, including accuracy, sensitivity, specificity, precision, recall, and F1 score, were calculated for each LLM-prompt-algorithm combination and presented as bar plots, with the value of each metric displayed vertically above the corresponding bars. Agreement in underlying reasoning was assessed by comparing the rationales used by biomedical LLMs and human experts to reach their conclusions. This was done by an independent assessor and the two human experts assessors. Disagreements were solved by consensus over plenary discussions. Results were visualized using a multi-panel plot: (i) bar plots showing question-level agreement and underlying reasoning agreement across algorithm questions, (ii) confusion matrices, (iii) bubble plots depicting inter-rater agreement, and (iv) summary bar charts of overall performance metrics. Inter-rater agreement was statistically evaluated using Gwet’s Agreement Coefficient 1 (AC1) [26]. Errors and inconsistencies were summarized and presented in tabular form and as heatmaps of frequency across questions of the Naranjo/WHO-UMC algorithms.

The R software environment was used for data analysis and, specifically, RStudio 2024.09.1+394 “Cranberry Hibiscus” (Build a1fe401fc08c232d470278d1bc362d05d79753d9, released on 2024-11-03) for Windows. To deploy the LLMs in a local Windows 11 environment, we used modular Python-based architecture (Python 3.10) that supports the inference of open-access transformer models. This architecture was built on a local server-client model, using Python libraries such as transformers, acceleration, and auto-gptq to ensure compatibility with a wide range of model formats, including GGUF and GPTQ. Model execution was handled via a backend using llama-cpp-python or text-generation-webui integrations, enabling seamless switching between quantified and full-precision variants. Graphics Processing Unit (GPU) acceleration was enabled where available through CUDA.

### 2.9 Reporting Standards

Methods and results were reported in accordance with the TRIPOD-LLM and CHART reporting standards (Supplementary Tables 3 and 4) [27,28]. In addition, the study adhered to the FAIR (Findable, Accessible, Interoperable, and Reusable) data principles to promote transparency and reproducibility [29]. All analysis code, prompts, and answers from LLMs as well as hyperparameter configurations are publicly available via a GitHub repository (Link: https://github.com/mauriziosessaku/Biomedical-LLMs-and-Prompt-Engineering-for-Causality-Assessment).

## 3. Results

ICSR demographics and clinical characteristics are reported in Table 2. A representative ICSR illustrating the assessments made by human experts and biomedical LLMs is shown in Supplementary Table 5.

**Table 2.** Individual Case Safety Report Characteristics.

|  | Level | Vaccine<br>(n=10) | High Prevalent<br>Drugs<br>(n=50) | Specialized<br>Drugs<br>(n=10) | High Risk<br>(n=10) | Rare Disease<br>(n=10) | Gene-Cell<br>Therapies<br>(n=25) | Newly<br>Approved Drugs<br>(n=35) | Total<br>(n=150) |
| --- | --- | --- | --- | --- | --- | --- | --- | --- | --- |
| **Age** | Median | 22 | 44 | 46.5 | 58 | 19 | 61.5 | 57.5 | 47 |
|  | (Range) | (14, 43) | (1, 82) | (7, 92) | (17, 89) | (0, 75) | (23, 87) | (21, 77) | (0, 92) |
|  | N/A | 3 | 19 | 4 | 7 | 4 | 13 | 15 | 65 |
| **Sex** | Male | 7 | 16 | 5 | 6 | 3 | 10 | 9 | 56 |
|  |  | (87.5) | (45.7) | (62.5) | (75.0) | (42.9) | (62.5) | (27.3) | (48.7) |
|  | Female | 1 | 19 | 3 | 2 | 4 | 6 | 24 | 59 |
|  |  | (12.5) | (54.3) | (37.5) | (25.0) | (57.1) | (37.5) | (72.7) | (51.3) |
|  | N/A | 2 | 15 | 2 | 2 | 3 | 9 | 2 | 34 |
| **Weight** | Median | - | 70 | 72 | - | 92.7 | 82 | 97.5 | 82 |
|  | (Range) |  | (3.4, 127) | (70, 95) |  | (92.7, 92.7) | (64, 165) | (90, 105) | (3.4, 165) |
|  | N/A | 10 | 38 | 7 | 10 | 9 | 20 | 33 | 127 |
| **Outcome** | Hospital | 10 | 14 | 4 | 4 | 2 | 4 | 2 | 40 |
|  |  | (100.0) | (36.8) | (44.4) | (44.4) | (28.6) | (21.1) | (33.3) | (40.8) |
|  | Disability | 0 | 2 | 0 | 1 | 0 | 0 | 0 | 3 |
|  |  | (0.0) | (5.3) | (0.0) | (11.1) | (0.0) | (0.0) | (0.0) | (3.1) |
|  | Other | 0 | 17 | 4 | 3 | 4 | 11 | 4 | 43 |
|  |  | (0.0) | (44.7) | (44.4) | (33.3) | (57.1) | (57.9) | (66.7) | (43.9) |
|  | Life-Threatening | 0 | 2 | 0 | 0 | 0 | 1 | 0 | 3 |
|  |  | (0.0) | (5.3) | (0.0) | (0.0) | (0.0) | (5.3) | (0.0) | (3.1) |
|  | Death | 0 | 2 | 1 | 1 | 1 | 3 | 0 | 8 |
|  |  | (0.0) | (5.3) | (11.1) | (11.1) | (14.3) | (15.8) | (0.0) | (8.2) |
|  | Congenital<br>Anomaly | 0 | 1 | 0 | 0 | 0 | 0 | 0 | 1 |
|  |  | (0.0) | (2.6) | (0.0) | (0.0) | (0.0) | (0.0) | (0.0) | (1.0) |
|  | N/A | 0 | 12 | 1 | 1 | 3 | 6 | 29 | 52 |
| **Number of**<br>**concomitant**<br>**drugs** | Median<br>(Range) | 1<br>(0, 6) | 3<br>(1, 101) | 2<br>(1, 5) | 4<br>(2, 14) | 1<br>(1, 7) | 1<br>(1, 14) | 1<br>(1, 2) | 1<br>(0, 101) |
| **Number of**<br>**concomitant**<br>**diseases** | Median<br>(Range) | 1<br>(0, 3) | 2<br>(1, 14) | 1.5<br>(1, 2) | 1.5<br>(1, 4) | 1<br>(1, 2) | 1<br>(1, 5) | 1<br>(1, 3) | 1<br>(0, 14) |
| **Number of**<br>**AEs** | Median<br>(Range) | 7.5<br>(4, 20) | 2<br>(1, 102) | 4.5<br>(1, 7) | 3<br>(1, 10) | 3<br>(1, 8) | 1<br>(1, 7) | 2<br>(1, 13) | 2<br>(1, 102) |

*Legend: AE = adverse events.*

### 3.1 Inter-rater agreement - TinyLlama 1.1B (Afrideva)-Naranjo-CoT

TinyLlama 1.1B (Afrideva)-Naranjo-CoT achieved >90% agreement for questions 4, 6-9 of the Naranjo algorithm. Agreement was moderate (44-62.7%) for questions on SmPC-listed AEs, temporal relationships, and objective evidence (Questions 1, 2, and 10), and lowest for alternative causes (Question 5). Overall adherence (56.7%) and the AC1 value (0.574) indicated moderate agreement, with most ICSRs classified as “Possible” by both experts and biomedical LLMs. Performance metrics reflected moderate accuracy and specificity, but low recall and sensitivity. Reasoning agreement was high for questions with clear factual anchors (Questions 4, 6-9), 82.7%, 83.3%, 89.3%, and 82%, respectively (Figure 1).

**Figure 1.**
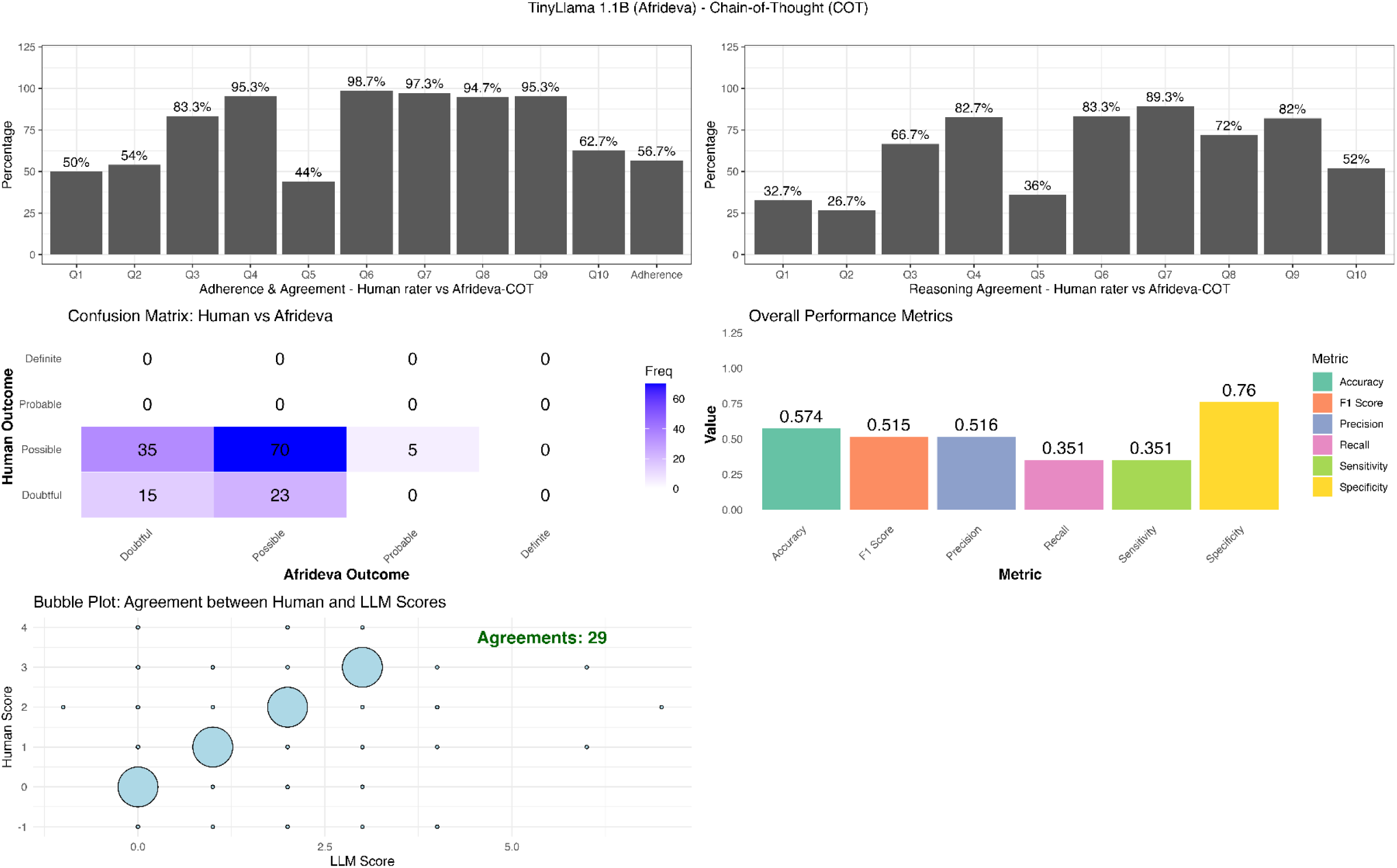
TinyLlama 1.1 B (Afrideva) - Naranjo - Chain-of-Thought Performance. Legend: a) Percentage of agreement for Q1 to Q10 of Naranjo Algorithm and overall Adherence between Human assessors and TinyLlama 1.1B (Afrideva)-CoT b) Percentage of reasoning agreement for Q1 to Q10 of Naranjo Algorithm and overall Adherence between Human assessors and TinyLlama 1.1B (Afrideva)-CoT c) Confusion Matrix plot for the 4 categories of classification “Doubtful”, “Possible”, “Probable”, “Definite” d) Metric plot for accuracy,F1, Precision, Recall, Specificity and Sensitivity e) Bubble plot for number of total agreement in the final numerical score of Naranjo between human assessors and TinyLlama 1.1B (Afrideva)-CoT. CoT =

### 3.2 Inter-rater agreement - TinyLlama 1.1B (Afrideva)-Naranjo-Decomposition

TinyLlama 1.1B (Afrideva)-Naranjo-Decomposition showed >80% agreement for questions 3-4 and 6-9 of the Naranjo algorithm. Agreement was lower (46.7-60.7%) for prior conclusive reports, temporal relationships, and objective evidence (Questions 1, 2, and 10), and lowest for prior conclusive reports (Question 1) and alternative causes (Question 5). Overall adherence (52.7%; AC1 = 0.547) also indicated moderate agreement, with most classifications falling into the “Possible” or “Doubtful” categories. Accuracy and specificity were moderate, whereas recall and sensitivity remained low. Reasoning agreement followed a similar pattern, being higher for structured, factual questions (Questions 3-4 and 6-9) and lower for those requiring clinical judgment (Questions 1, 2, and 5) (Figure 2).

**Figure 2.**
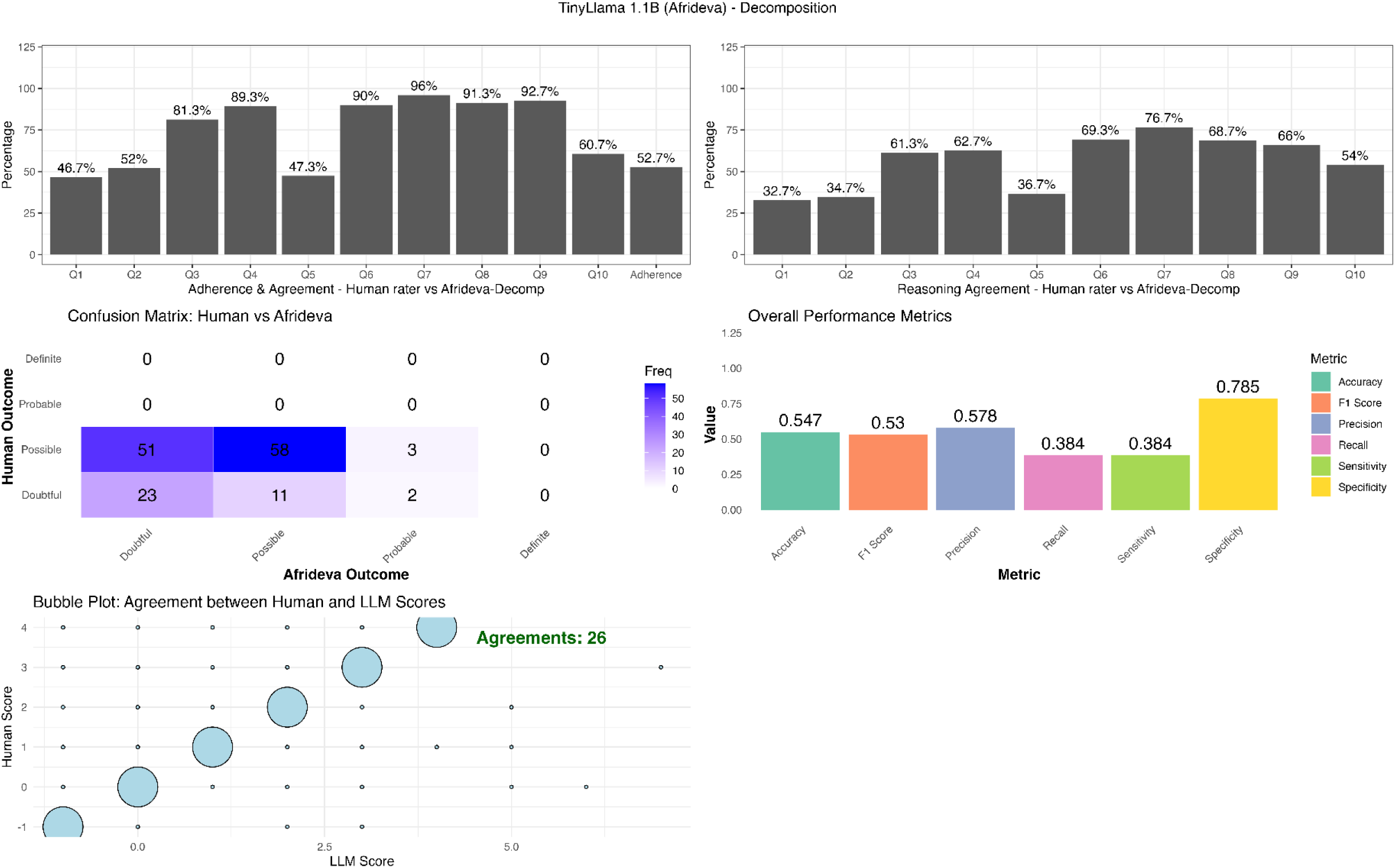
TinyLlama 1.1 B (Afrideva) - Naranjo - Decomposition Performance. Legend: a) Percentage of agreement for Q1 to Q10 of Naranjo Algorithm and overall Adherence between Human assessors and TinyLlama 1.1B (Afrideva)-Decomposition b) Percentage of reasoning agreement for Q1 to Q10 of Naranjo Algorithm and overall Adherence between Human assessors and TinyLlama 1.1B (Afrideva)-Decomposition c) Confusion Matrix plot for the 4 categories of classification “Doubtful”, “Possible”, “Probable”, “Definite” d) Metric plot for accuracy, F1, Precision, Recall, Specificity and Sensitivity e) Bubble plot for number of total agreement in the final numerical score of Naranjo between human assessors and TinyLlama 1.1B (Afrideva)-Decomposition. <u>Note</u>: 2 cases were not assessable by the LLM.

### 3.3 Inter-rater agreement - Medicine Llama3 8B (QuantFactory)-Naranjo-CoT

Medicine Llama3 8B (QuantFactory)-Naranjo-CoT yielded the strongest alignment, with >80% agreement for questions 3-4 and 6-9 of the Naranjo algorithm. Agreement for temporal relationships, alternative causes, and objective evidence (Questions 2, 5, and 10) ranged from 52.7% to 63.7%, whereas Question 1 showed the lowest agreement. Overall adherence was highest among the three configurations (63.3%; AC1 = 0.644), indicating strong agreement, with the majority of ICSRs classified as “Possible” (n = 96). Accuracy and specificity were moderate, but, as with the other models, recall and sensitivity remained low. Reasoning agreement was highest for Questions 3, 4, 6, 7, 8, and 9, 73.3%, 82.7%, 92%, 82%, 74.7%, and 77.3%, respectively (Figure 3).

**Figure 3.**
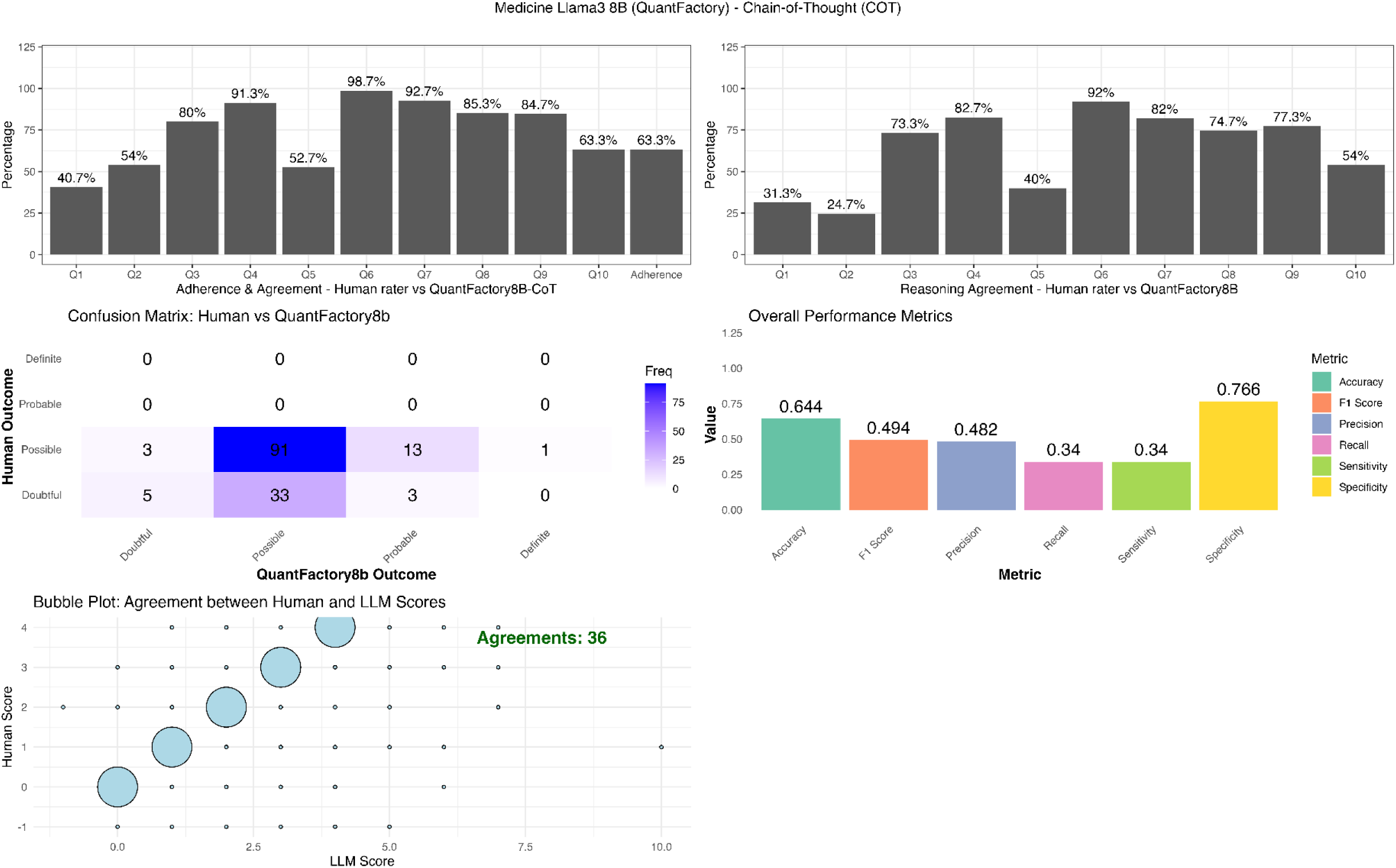
Medicine Llama3 8B (QuantFactory) - Naranjo - Chain-of-Thought Performance. Legend: a) Percentage of agreement for Q1 to Q10 of Naranjo Algorithm and overall Adherence between Human assessors and Medicine Llama3 8B (QuantFactory)-CoT b) Percentage of reasoning agreement for Q1 to Q10 of Naranjo Algorithm and overall Adherence between Human assessors and Medicine Llama3 8B (QuantFactory)-CoT c) Confusion Matrix plot for the 4 categories of classification “Doubtful”, “Possible”, “Probable”, “Definite” d) Metric plot for accuracy,F1, Precision, Recall, Specificity and Sensitivity e) Bubble plot for number of total agreement in the final numerical score of Naranjo between human assessors and Medicine Llama3 8B (QuantFactory)-CoT. CoT = Chain-of-Thought. <u>Note</u>: 1 case was not assessable by the LLM.

### 3.4 Inter-rater agreement - Medicine Llama3 8B (QuantFactory)-WHO-UMC-CoT

The Medicine Llama3 8B (QuantFactory)-WHO-UMC-CoT combination exceeded 80% agreement only for Question 4 (strong evidence against causal association). Agreement for strong evidence for other causes (Question 1), known causal association with the drug/vaccine (Question 2), time window of increased risk (Question 3), and other qualifying factors (Question 5) ranged from 46.7% to 54.7%. Overall adherence was 23.3%, with an AC1 of 0.297, indicating minimal agreement on final causality classification. Although classifications were distributed across categories, most fell within the “Possible” category for both evaluators. Performance metrics indicated moderate accuracy (0.297) and specificity (0.933), while recall (0.18) and sensitivity (0.188) were lower. Precision and F1 score were 0.2 and 0.192, respectively. Reasoning agreement was poor, except for Question 4 (71.3%) (Supplementary Figure 1).

### 3.5 Inter-rater agreement - Medicine Llama3 8B (QuantFactory)-WHO-UMC-Decomposition

Medicine Llama3 8B (QuantFactory)-WHO-UMC-Decomposition showed limited performance, with >76% agreement for questions related to strong evidence against causal association (Question 4) and 51.3-76.7% agreement for other causes, known causal association with the drug/vaccine, and time window of increased risk (Questions 1-3). Overall adherence remained low (24.7%; AC1 = 0.327), indicating a minimal level of overall agreement. Although most ICSRs were classified as “Possible,” performance metrics indicated moderate accuracy (0.84) and low specificity (0.233). Recall (0.233) and sensitivity (0.233) were also low. Precision and F1 score were 0.253 and 0.587, respectively. Reasoning agreement was extremely low across all WHO questions (Questions 1-5) (Supplementary Figure 2).

### 3.6. Head-to-Head Comparisons

Combinations of biomedical LLMs and prompt strategies using the Naranjo algorithm (i.e., TinyLlama 1.1B (Afrideva)-CoT, TinyLlama 1.1B (Afrideva)-Decomposition, and Medicine LLaMA-3 8B (QuantFactory)-CoT) followed a similar performance pattern (Supplementary Figure 3). In contrast, the two biomedical LLM-prompt combinations using the WHO-UMC algorithm showed distinct performance patterns. Agreement for WHO-UMC Questions 1 and 3 was similar across both models, whereas larger differences emerged for Questions 2, 4, 5, and 6.

MedLLaMA v20 (JL42)-Decomposition achieved higher agreement for WHO-UMC Question 2 and for the final causality classification, while Medicine LLaMA-3 8B (QuantFactory)-CoT performed better for Questions 4 and 5. MedLLaMA v20 (JL42)-Decomposition showed minimal reasoning alignment, with values ranging from 1% to 3% across all WHO-UMC questions. In contrast, Medicine LLaMA-3 8B (QuantFactory)-CoT demonstrated substantially higher reasoning agreement, with median values around 40% for most questions and a peak of 71.3% for Question 4, representing the highest reasoning alignment observed between LLMs and human assessors within the WHO-UMC framework (Supplementary Figure 4).

### 3.7. Errors and Inconsistencies

The TinyLlama 1.1B (Afrideva)-Naranjo-CoT combination showed substantial variability, with error-free responses ranging from 40% to 70% across Questions 1-10. Performance improved with the TinyLlama 1.1B (Afrideva)-Naranjo-Decomposition strategy, which achieved error-free rates between 52% and 65%. In contrast, the Medicine LLaMA-3 8B (QuantFactory)-Naranjo-CoT strategy demonstrated the most consistent and robust performance, with 72%-74% of cases free of errors across all questions and an 87% error-free rate for the final causality score.

Within the WHO framework, the Medicine LLaMA-3 8B (QuantFactory)-WHO-CoT combination showed stable accuracy (60%-71%) and a 72% error-free final score. By comparison, the MedLLaMA v20 (JL32)-WHO-Decomposition configuration exhibited the poorest performance, with only 2%-4% of cases free of procedural errors, reflecting substantial inconsistencies across all questions (Supplementary Figure 5).

Error pattern analysis revealed distinct failure modes across configurations. For both TinyLlama 1.1B (Afrideva) Naranjo-based strategies, the most frequent issues were instruction drift. Errors were less frequent for Medicine LLaMA-3 8B (QuantFactory)-Naranjo-CoT; however, prompt echoing and lack of justification were the most common categories. Despite these errors, this configuration exhibited the lowest overall error rate among the five strategies (Supplementary Figure 6).

In contrast, the Medicine LLaMA-3 8B (QuantFactory)-WHO-Decomposition combination frequently produced looping self-dialogue and chat-like drafts, suggesting difficulty maintaining structured reasoning under the WHO framework. The MedLLaMA v20 (JL42)-WHO-Decomposition combination showed the highest overall error burden, with lack of justification observed in nearly 147 of 150 ICSRs (Supplementary Figure 7).

Representative examples of inconsistencies and errors across all five combinations, along with the corresponding case identifiers, are provided in Supplementary Table 6.

## 4. Discussion

This study assessed the performance of biomedical LLMs, combined with prompt engineering and different causality algorithms, for causality assessment in ICSRs. The combinations of LLM-prompt-algorithm were tested in a heterogenous dataset comprised of 150 real-world ICSRs spanning diverse drug classes and COVID-19 vaccine reports [30].

### 4.1 Training Sources

Across prompt-algorithm pairings, biomedical LLMs showed improved performance over general-purpose LLMs tested by Abate et al. [8], reaching up to 64% inter-rater agreement with human experts, almost doubling the alignment previously reported for expert causality tasks (34%) [8]. A possible reason for this increased performance could be the training sources of the LLMs. General-purpose LLMs primarily learn from open-web or web-crawled sources [31]. Although general-purpose LLMs may encode a wider linguistic spectrum, they lack the domain-focused knowledge that is needed to capture the reasoning logic behind AE/AEFI temporal and biological plausibility, alternative causes, and objective evidence [32]. Biomedical LLMs instead, are trained on biomedical literature and therefore less exposed to misinformation or over-general linguistic associations [33,34]. When the training sources are based on biomedical literature, the biomedical LLM seems to better approximate human clinical reasoning, which is crucial in causality assessment [35].

### 4.2 Model Size, Explainability, and Human-in-the-loop

It is important to highlight that biomedical LLMs tested in this study had different sizes (i.e., number of parameters) but they all were smaller than general purpose LLMs tested by Abate et al.,. As it has been widely described in the scientific literature, model size does not guarantee better causal reasoning and may, paradoxically, yield highly confident but unverified or inaccurate statements [36]. Intuitively, it may be expected that reducing the size of the LLM and training it on biomedical literature would overcome the aforementioned problems. However, in this study, we found that such practices did not improve performance in crucial aspects of the causality assessment process, such as the assessment of temporal and biological plausibility, alternative causes, and objective evidence. This result is not surprising, as noted by Dada et al. in other contexts [37]. Specifically, Dada et al. have found that optimizing domain knowledge can inadvertently degrade instruction-following or increase hallucination tendencies and procedural errors unless additional alignment techniques, such as instruction-tuning or model merging, remain part of the optimization process [37]. Instruction-drift errors were also observed, particularly when prompts contained multiple constraints or complex reasoning steps, leading to output that partially deviated from the instructions of the algorithm. In this study, by analyzing the errors and inconsistencies of the biomedical LLMs, in agreement with Dada et al. [37], we have noticed that the gain in clinical reasoning obtained by using biomedical literature as training source did not outweigh the risk of misunderstanding the causality assessment algorithm instructions in the prompt and generating viable output. Specifically, we have seen that on multiple occasions even the best-performing biomedical LLM-prompt-algorithm combinations had recurring error patterns that would constrain their usability in causality assessment. One notable behavior was the tendency to syntactically mirror parts of the input prompt in the generated answer, a pattern previously described as prompt-echoing [38], which suggests that correct scores may at times derive from linguistic mimicry rather than independent reasoning. Therefore, even if the biomedical LLM reached a good inter-rater agreement for questions’ scoring, the inability to correctly reason and support with evidence the provided score undermines their usefulness. Especially in settings (e.g., European Union) where there is a need of having a human-in-the-loop to take the final decision, an undocumented score may not be useful, as it will not provide the human with the necessary information to take an informed decision, which compromises transparency and traceability, as well as explainability, that are core requirements for implementation of these tools in routine pharmacovigilance [39].

### 4.3 Epistemic Uncertainty

We noted that biomedical LLMs showed limited sensitivity to epistemic uncertainty. Even when ICSRs contained incomplete or ambiguous evidence, human experts frequently opted for guarded judgement or assigned “unknown” values, whereas the biomedical LLMs tended to generate high-certainty categorical responses regardless of missing information, potentially creating unreliable causal assertions.

### 4.4 Role of the Algorithm and Prompt

In this study, we noticed that the performance of the biomedical LLM was strongly modulated by choice of the causality assessment algorithm. When LLM prompting and model choice were kept constant, causal classification agreement still shifted dramatically depending on the causality algorithm used. The combination of Medicine LLaMA-3 8B (QuantFactory) with a Chain-of-Thought prompt achieved a 63.3% exact-label agreement with humans when responses were structured under the Naranjo algorithm, but the same model-prompt pair dropped to 23.3% agreement when applied to the WHO-UMC approach. These results suggest that compatibility between LLMs and causality tools is not a secondary technical detail but a foundational design requirement. Even when biomedical knowledge improves clinical reasoning, some established human-centric causality frameworks may not seamlessly translate into LLM decision logic.

Another important insight from our study is that prompt strategy had little impact on the performance. When the same biomedical LLM and the Naranjo algorithm were paired with either Chain-of-Thought or Decomposition, inter-rater agreement appeared similar. This could be misread as evidence that prompting has minimal influence, yet a broader evaluation across multiple biomedical and non-biomedical models demonstrated the opposite [40]. Although a key benefit of the prompt strategy is to promote reasoning by partitioning the task into discrete steps, this structural advantage is neutralized when the causality algorithm itself dictates a stepwise approach to causality assessment, ultimately causing the prompts to converge to a similar structure and, therefore, performance. When examining algorithmic fidelity, the picture becomes even clearer. The best-performing configuration, combining Medicine LLaMA-3 8B (QuantFactory) with the Chain-of-Thought prompting and the Naranjo algorithm, produced an exact numerical score match in only 36 of 150 ICSRs. While this might initially appear to reflect limited capacity to reproduce the incremental logic of the tool, a more granular interpretation shows that score deviations were typically minor, usually 1–2 points from the expert-assigned score, and almost never shifted the final causal classification (e.g., Possible, Probable). This indicates that the model is often capturing the semantic weighting and internal decision structure of item-level causality correctly despite imperfect numeric replication, and that these small numeric differences rarely introduce meaningful classification disagreement in ICSR interpretations. The findings therefore suggest that itemized causality tools such as Naranjo maintain strong categorical robustness even when adopted by machine reasoning systems, while narrative-dependent frameworks have repeatedly shown lower performance.

### 4.5 Elements of the Causality Assessment for which the Biomedical LLMs Failed

Deeper analysis of how biomedical LLMs responded to individual items in AE/AEFI causality assessment shows that performance limitations are not uniform across all elements. Within the Naranjo algorithm, Questions 1 (Listedness), 2 (Temporal plausibility), 5 (Alternative causes), and 10 (Objective Evidence) repeatedly emerged as sources of errors. These questions require semantic alignment, temporal reconstruction, and differentiation between different types clinical data and ICSR elements, reasoning steps that are performed implicitly in human assessment but remain difficult for current biomedical LLMs to resolve consistently.

For Question 1, which asks whether the AE/AEFI is listed in the official product information (i.e., SmPC) or in the scientific literature, we believe that it was to be expected to have a low performance of biomedically trained LLMs. Training sources of biomedical LLMs may not include the SmPCs, therefore it may be unfair to expect that biomedical LLMs were able to perform the task. We also believe that perhaps more sophisticated tools like AI agents or Agentic AI, which have access to tools for different tasks (e.g., accessing the web and, therefore, the latest available SmPC) may lead to a better performance in the future.

### 4.6. Strengths and Limitations

The robustness of this investigation is grounded in several key methodological advantages, beginning with the implementation of a regulatory-grade gold standard for performance comparison. By utilizing high-quality causality assessments conducted by expert human assessors, specifically a senior medical doctor from a pharmaceutical company and a specialist pharmacist, this study ensured that the reference standard met rigorous industry and regulatory expectations. Furthermore, the evaluation was performed on a heterogeneous dataset of 150 real-world ICSRs rather than focusing on a single drug class. This diverse sample spanned several distinct categories, including newly approved drugs, gene and cell therapies, commonly prescribed medications, and clinically confirmed vaccine reports from the VAERS database.

The study’s comparative framework is a further strength, as it tested two internationally recognized causality assessment algorithms with divergent structural characteristics: the itemized Naranjo Adverse Drug Reaction Probability Scale and the narrative-dependent WHO-UMC algorithm. This was complemented by a systematic evaluation of multiple biomedical-specific LLMs across various state-of-the-art prompt engineering strategies such as Chain-of-Thought and Decomposition. Finally, the commitment to high accessibility and reproducibility is evidenced using open-source models and adherence to FAIR principles, with all analysis code and prompt templates made publicly available to facilitate future academic and clinical feasibility evaluations.

Despite these methodological strengths, several limitations must be acknowledged. A primary constraint was the lack of systematic hyperparameter tuning, as model performance was assessed using off-the-shelf configurations; optimizing these parameters represents a critical next methodological step. Additionally, the reliance on a single-model decision pipeline may have limited performance. Future research utilizing “Agentic AI” to route specific subtasks to specialized models could potentially overcome current limitations regarding instruction stability and reasoning. The inherent “noise” and “poor data quality” within spontaneous reporting databases like FAERS and VAERS also introduces challenges, as missing clinical evidence, inconsistent documentation, and a lack of onset chronology can hinder both human and machine reasoning. Technical and reasoning constraints were also observed, including “frozen” knowledge cutoffs and small parameter scales, which contributed to errors such as prompt echoing, instruction drift, and an inconsistent ability to provide objective justification.

## 5. Conclusions

The findings of this study demonstrate that biomedical LLMs can approximate structured causality assessment for ICSRs more reliably than the general-purpose models previously evaluated in the literature. By utilizing domain-specific training data, these models achieved nearly double the alignment with human experts compared to general-purpose models. Algorithm compatibility emerged as the primary determinant of alignment between human and biomedical LLM assessments. While itemized frameworks like the Naranjo algorithm demonstrated strong categorical robustness, narrative-dependent structures, such as the WHO-UMC approach, revealed significant interpretation gaps from biomedical LLMs. Furthermore, the persistent variability in interpreting critical concepts, such as “objective evidence” and “alternative causes”, underscores the necessity for closer alignment between LLM outputs and established regulatory expectations. Despite the improved performance of biomedical LLMs, a lack of consistent reasoning and proper substantiation of scores still undermines their current usability in highly regulated settings such as Pharmacovigilance. In individual-level causality assessment, where “human-in-the-loop” workflows are required (i.e., Europe), expert feedback and final decisions must be based on transparent and explainable outcomes. Therefore, further research is necessary to identify the optimal LLM configurations to support decision-makers in pharmacovigilance. As these LLMs evolve through refined training sources, prompting, systematic hyperparameter tuning, and more advanced orchestration, such as Agentic AI, their integration into signal management processes holds the potential to significantly enhance the consistency, efficiency, and reproducibility of safety monitoring.

## Data Availability

All data produced in the present work are contained in the manuscript

## 6. Ethics, Funding, & Conflict of Interest

No funding was received for this study. Ethical approval and informed consent were not required for this study as it utilized exclusively open-source, anonymized data. The authors declare that there are no conflicts of interest regarding the conduct or reporting of this study.

## 7. Authors’ contribution

NSH and MS were responsible for the substantial conception and design of the study, as well as the acquisition, analysis, and interpretation of data and the drafting of the manuscript. DGP contributed significantly to the acquisition, analysis, and interpretation of data and participated in drafting the work. MAB and VB provided substantial contributions to the design of the work and the critical revision of the manuscript for important intellectual content. SNM, SOS, and LM contributed to the acquisition of data and ongoing inputs on the design of the study, and with SNM specifically serving as a human expert assessor for the gold-standard reference, and all three authors critically revised the manuscript. MS provided final approval of the version to be published and served as the corresponding author. All authors agree to be accountable for all aspects of the work in ensuring that questions related to the accuracy or integrity of any part of the study are appropriately investigated and resolved.

