## Supplementary material for "Biomedical Large Language Models and Prompt Engineering for Causality Assessment of Individual Case Safety Reports in Pharmacovigilance"

[(4)]Safety Operations, Novo Nordisk, Søborg, Denmark.

**Corresponding author:**

Maurizio Sessa,

Department of Drug Design and Pharmacology,

University of Copenhagen,

Jagtvej 160, Copenhagen 2100, Denmark

**Supplementary Table 1**. FAERS and VAERS ICSR Structured Template.

| **FAERS VARIABLES** |
|---|
| CASE ID |
| CASE VERSION |
| EVENT DATE |
| AUTHORIZATION NUMBER |
| MANUFACTURER NUMBER |
| MANUFACTURER SENDER |
| LITERATURE REFERENCE |
| AGE |
| AGE CODE |
| AGE GROUP |
| SEX |
| E SUB |
| WEIGHT |
| WEIGHT CODE |
| OCCUPATION CODE |
| REPORTER COUNTRY |
| OCCURENCE COUNTRY |
| ADVERSE REACTION(S) |
| CASE ID NUMBER |
| DRUG SEQUENCE |
| ROLE CODE |
| DRUG NAME |
| PRODUCT AI |
| VAL VBM |
| ROUTE |
| DOSE VBM |
| CUMULATIVEDOSE (CHAR) |
| CUMULATIVE DOSE (UNIT) |
| DECHALLANGE |
| REACHALLANGE |
| LOT NUMBER |
| EXPIRATION DATE |
| NDA NUMBER |
| DOSE AMOUNT |

| |
|---|
| DOSE UNIT |
| DOSE FORM |
| DOSE FREQUENCY |
| OUTCOME CODE |
| REPORT SOURCE CODE |
| INDICATION FOR USE |
| THERAPY START DATE |
| THERAPY END DATE |
| DURATION |
| DURATION CODE |

*Legend: NA = Not Announced, CN = Consumer, YR = Year, FAERS = Food and Drug Administration Adverse Event Reporting System*

| **VAERS VARIABLES** |
|---|
| VAERS_ID |
| S |
| M |
| H |
| V |
| D |
| O |

*Legend: VAERS = Vaccine Adverse Event Reporting System, ID = Identity, S = symptoms/scenario the person has experienced after the vaccine, M = other medication the person is taking that may interact with the vaccine, H = previous conditions/disease the person had and laboratory data, V = vaccine administered to the person, D = number of days between the vaccine administration and onset of the symptoms, O = outcome and symptoms as a potential effect of the vaccine that has to be investigated.*

**Supplementary Table 2.** Template for Data Collection.

| V1 | V2 | V3 | V4 | V5 | V6 | V7 | V8 | V9 | V1<br>0 | … | V35 | V3<br>6 | V37 | V38 | V3<br>9 | V40 | … | V88 |
|---|---|---|---|---|---|---|---|---|---|---|---|---|---|---|---|---|---|---|

Columns are labeled V1-V71 to allow compact visualization on a single horizontal page. Detailed variable definitions, formats, and allowed values are provided below.

| Variable | Description | Type | Format |
|---|---|---|---|
| V1 | Case number (1 to 150) | Integer | Numeric |
| V2 | FAERS/VAERS case identifier | Integer | Numeric |
| V3 | Number of Naranjo questions with no LLM score | Integer | Numeric |
| V4 | Total number of Naranjo questions evaluated by the LLM | Integer | Numeric |
| V5–-V34 | Q1–Q10 (Naranjo): human score, LLM score, and agreement (1 for yes, 0 for no) | Mixed | Numeric / Binary |
| V35 | Total score for Naranjo (human) | Integer | Numeric |
| V36 | Total score for Naranjo (LLM) | Integer | Numeric |
| V37 | Final Naranjo causality adherence (human vs LLM) | Binary | 0 / 1 |
| V38-53 | Q1–Q5 (WHO-UMC): human score, LLM score, and agreement (1 for yes, 0 for no) | Mixed | Text / Binary |
| V54 | WHO-UMC classification (human) | Categorical | Text |
| V55 | WHO-UMC classification (LLM) | Categorical | Text |
| V56 | WHO-UMC adherence (human vs LLM) | Binary | 0 / 1 |
| V57-V77 | Q1–Q10 (Naranjo) reasoning text and agreement | Mixed | Text / Binary |
| V78-V88 | Q1-Q5 (WHO-UMC) reasoning text and agreement | Mixed | Text / Binary |

**Supplementary Table 3.** TRIPOD-LLM Checklist.

| Section | Item | Description | Page no. |
|---|---|---|---|
| **Title** | 1 | Identify the study as developing, fine-tuning and/or evaluating the performance of an LLM, specifying the task, the target population and the outcome to be predicted. | 1 |
| **Introduction** | | | |
| **Abstract** | 2 | See TRIPOD-LLM for abstracts. | 2 |
| **Background** | 3a | Explain the healthcare context/use case (for example, administrative, diagnostic, therapeutic and clinical workflow) and rationale for developing or evaluating the LLM, including references to existing approaches and models. | 3 |
| | 3b | Describe the target population and the intended use of the LLM in the context of the care pathway, including its intended users in current gold standard practices (for example, healthcare professionals, patients, public or administrators). | 3 |
| **Objectives** | 4 | Specify the study objectives, including whether the study describes the initial development, fine-tuning or validation of an LLM (or multiple stages). | 3 |
| **Methods** | | | |
| **Data** | 5a | Describe the sources of data separately for the training, tuning and/or evaluation datasets and the rationale for using these data (for example, web corpora, clinical research/trial data, EHR data or unknown). | 3-4 |
| | 5b | Describe the relevant data points and provide a quantitative and qualitative description of their distribution and other relevant descriptors of the dataset (for example, source, languages and countries of origin). | 3-4 |
| | 5c | Specifically state the date of the oldest and newest item of text used in the development process (training, fine-tuning and reward modeling) and the evaluation datasets. | N/A |
| | 5d | Describe any data preprocessing and quality checking, including whether this was similar across text corpora, institutions and relevant sociodemographic groups. | 3-4 |
| | 5e | Describe how missing and imbalanced data were handled and provide reasons for omitting any data. | N/A |
| **Analytical Methods** | 6a | Report the LLM name, version and last date of training. | 5 |
| | 6b | Report details of the LLM development process, such as LLM architecture, training, fine-tuning procedures and alignment strategy (for example, reinforcement learning and direct preference optimization) and alignment goals (for example, helpfulness, honesty and harmlessness). | 5 |
| | 6c | Report details of how the text was generated using the LLM, including any prompt engineering (including consistency of outputs), and inference settings (for example, seed, temperature, max token length and penalties), as relevant. | 5-6 |
| | 6d | Specify the initial and postprocessed output of the LLM (for example, probabilities, classification and unstructured text). | 5-6 |
| | 6e | Provide details and rationale for any classification and, if applicable, how the probabilities were determined and thresholds identified. | 6-7 |
| **LLM Output** | 7a | Include metrics that capture the quality of generative outputs, such as consistency, relevance, accuracy and presence/type of errors compared to gold standards. | 4-7 |
| | 7b | Report the outcome metrics' relevance to the downstream task at deployment time and, where applicable, the correlation of metric to human evaluation of the text for the intended use. | 6-7 |
| | 7c | Clearly define the outcome, how the LLM predictions were calculated (for example, formula, code, object and API), the date of inference for closed-source LLMs and evaluation metrics. | In the GitHub |

| | | | |
|---|---|---|---|
| | 7d | If outcome assessment requires subjective interpretation, describe the qualifications of the assessors, any instructions provided, relevant information on demographics of the assessors and inter-assessor agreement. | 4-5 |
| | 7e | Specify how performance was compared to other LLMs, humans and other benchmarks or standards. | 6-7 |
| **Annotation** | 8a | If annotation was done, report how the text was labeled, including providing specific annotation guidelines with examples. | N/A |
| | 8b | If annotation was done, report how many annotators labeled the dataset(s), including the proportion of data in each dataset that was annotated by more than one annotator, and the inter-annotator agreement. | N/A |
| | 8c | If annotation was done, provide information on the background and experience of the annotators or the characteristics of any models involved in labeling. | N/A |
| **Prompting** | 9a | If research involved prompting LLMs, provide details on the processes used during prompt design, curation and selection. | 5-6 |
| | 9b | If research involved prompting LLMs, report what data were used to develop the prompts. | 5-6 |
| **Summarization** | 10 | Describe any preprocessing of the data before summarization. | N/A |
| **Instruction tuning/ alignment** | 11 | If instruction tuning/alignment strategies were used, what were the instructions, data and interface used for evaluation, and what were the characteristics of the populations doing the evaluation? | N/A |
| **Compute** | 12 | Report compute, or proxies thereof (for example, time on what and how many machines, cost on what and how many machines, inference time, floating-point operations per second), required to carry out methods. | N/A |
| **Ethical approval** | 13 | Name the institutional research board or ethics committee that approved the study and describe the participant-informed consent or the ethics committee waiver of informed consent. | 15 |
| **Open science** | 14a | Give the source of funding and the role of the funders for the present study. | 15 |
| | 14b | Declare any conflicts of interest and financial disclosures for all authors. | 15 |
| | 14c | Indicate where the study protocol can be accessed or state that a protocol was not prepared. | N/A |
| | 14d | Provide registration information for the study, including register name and registration number, or state that the study was not registered. | N/A |
| | 14e | Provide details of the availability of the study data. | 7 |
| | 14f | Provide details of the availability of the code to reproduce the study results. | 7 |
| **Public involvement** | 15 | Provide details of any patient and public involvement during the design, conduct, reporting, interpretation or dissemination of the study or state no involvement. | N/A |
| **Results** | | | |
| **Participants** | 16a | When using patient/EHR data, describe the flow of text/EHR/patient data through the study, including the number of documents/questions/participants with and without the outcome/label and follow-up time as applicable. | 3-4 |
| | 16b | When using patient/EHR data, report the characteristics overall and for each data source or setting and development/evaluation splits, including the key dates, key characteristics and sample size. | 7 |
| | 16c | For LLM evaluation that includes clinical outcomes, show a comparison of the distribution of important clinical variables that may be associated with the outcome between development and evaluation data, if available. | N/A |
| | 16d | When using patient/EHR data, specify the number of participants and outcome events in each analysis (for example, for LLM development, hyperparameter tuning and LLM evaluation). | N/A |
| **Performance** | 17 | Report LLM performance according to prespecified metrics (see item 7a) and/or | 7-10 |

| | | | |
|---|---|---|---|
| | | human evaluation (see item 7d). | |
| **LLM updating** | 18 | If applicable, report the results from any LLM updating, including the updated LLM and subsequent performance. | N/A |
| **Discussion** | | | |
| **Interpretation** | 19a | Give an overall interpretation of the main results, including issues of fairness in the context of the objectives and previous studies. | 10-14 |
| **Limitations** | 19b | Discuss any limitations of the study and their effects on any biases, statistical uncertainty and generalizability. | 10-14 |
| **Usability of the LLM in context** | 19c | Describe any known challenges in using data for the specified task and domain context with reference to representation, missingness, harmonization and bias. | 10-14 |
| | 19d | Define the intended use for the implementation under evaluation, including the intended input, end-user and level of autonomy/human oversight. | 10-14 |
| | 19e | If applicable, describe how poor quality or unavailable input data should be assessed and handled when implementing the LLM; that is, what is the usability of the LLM in the context of current clinical care. | 10-14 |
| | 19f | If applicable, specify whether users will be required to interact in the handling of the input data or use of the LLM, and what level of expertise is required of users. | 10-14 |
| | 19g | Discuss any next steps for future research, with a specific view of the applicability and generalizability of the LLM. | 10-14 |

*Legend: M = LLM methods; D = de novo LLM development; E = LLM evaluation; H = LLM evaluation in healthcare settings; C = classification; OF = outcome forecasting; QA = long-form question answering; IR = information retrieval; DG = document generation; SS = summarization and simplification; MT = machine translation; API = application programming interface.*

**Supplementary Table 4.** CHART Checklist.

| Heading | No | Chart checklist item | Page no. |
|---|---|---|---|
| **Title and Abstract** | | | |
| **Title** | 1a | State that the study is assessing one or more generative AI-driven chatbots for clinical evidence or health advice. | 1 |
| **Abstract/ Summary** | 1b | Apply a structured format, if applicable. | 2 |
| **Introduction** | | | |
| **Background** | 2a | State the scientific background, rationale, and healthcare context for evaluating the generative AI-driven chatbot(s), referencing relevant literature when applicable. | 3 |
| | 2b | State the aims and research questions including the target audience, intervention, comparator(s), and outcome(s). | 3 |
| **Methods** | | | |
| **Model Identifiers** | 3a | State the name and version identifier(s) of the generative AI model(s) and chatbot(s) under evaluation, as well as their date of release or last update. | 5 |
| | 3b | State whether the generative AI model(s) and chatbot(s) are open-source or closed-source/proprietary. | 5 |
| **Model Details** | 4a | State whether the generative AI model was a base model or a novel base model, tuned model, or fine-tuned model. | 5 |
| | 4b | If a base model is used, cite its development in sufficient detail to identify the model. | 5 |
| | 4c | If a novel base model, tuned model, or fine-tuned model is used, describe the pre- and/or post-implementation/deployment data and parameters. | N/A |
| **Prompt Engineering** | 5a | Describe the evolution of study prompt development. | 5-6 |
| | 5ai | Describe the sources of prompts. | 5-6 |
| | 5aii | State the number and characteristics of the individual(s) involved in prompt engineering. | 5-6 |
| | 5aiii | Provide details of any patient and public involvement during prompt engineering. | N/A |
| | 5b | Provide study prompts. | 5-6 |
| **Query Strategy** | 6a | State route of access to generative AI model. | 5 |
| | 6b | State the date(s) and location(s) of queries for the generative AI-driven chatbot(s) including the day, month, and year as well as city and country. | 5 |
| | 6c | Describe whether prompts were input into separate chat session(s). | 5 |
| | 6d | Provide all generative AI-driven chatbot output/responses | Available on GitHub |
| **Prompting** | 7a | Define the ground truth or reference standard used to define successful generative AI-driven chatbot performance. | 4-5 |
| | 7b | Describe the process undertaken for generative AI-driven chatbot performance evaluation. | 4-5 |
| | 7bi | State the number and characteristics of team members involved in performance evaluation. | 4-5 |
| | 7bii | Provide details of any patients and public involvement during the evaluation process. | N/A |
| | 7biii | State whether evaluators were blinded to the identity of the generative AI-driven chatbot(s) under assessment. | N/A |
| **Sample Size** | 8 | Report how the sample size was determined. | N/A |
| **Data Analysis** | 9a | Describe statistical analysis methods, including any evaluation of reproducibility of generative AI-driven chatbot responses. | 6-7 |
| | 9ai | Report the measures used for performance evaluation. | 6-7 |

| | | | |
|---|---|---|---|
| **Results** | | | |
| | 10a | Report the performance evaluation undertaken including the alignment between generative AI-driven chatbot output and ground truth or reference standard using quantitative or mixed methods approaches as applicable. | 7-10 |
| | 10b | For responses deviating from the ground truth or reference standard, state the nature of the difference(s). | 7-10 |
| | 10c | Indicate where the study protocol can be accessed or state that a protocol was not prepared. | N/A |
| **Discussion** | | | |
| | 11a | Interpret study findings in the context of relevant evidence. | 10-14 |
| | 11b | Describe the strengths and limitations of the study. | 10-14 |
| | 11c | Describe the potential implications for practice, education, policy, regulation, and research. | 10-14 |
| **Open Science** | | | |
| **Disclosures** | 12a | Report any relevant conflicts of interest for all authors. | 15 |
| **Funding** | 12b | Report sources of funding and their role in the conduct and reporting of the study. | 15 |
| **Ethics** | 12c | Describe the process undertaken for ethical approval. | 15 |
| | 12ci | Describe the measures taken to safeguard data privacy of patient health information, as applicable. | 15 |
| | 12cii | State whether permission/licensing was obtained for the use of original, copyrighted data. | 15 |
| **Protocol** | 12d | Provide a study protocol. | N/A |
| **Data availability** | 12e | State where study data, code repository, and model parameters can be accessed. | 7 |

**Supplementary Table 5.** Human Expert and LLM Assessment of Example Case.

<table>
<tr><td colspan="2"><u>Case description</u><br>Case ID: 242444272 ; Event Date: OMITTED ; FDA Date: OMITTED ; Manufacturer Sender: NOVARTIS ; Literature Reference: NA ; Age: OMITTED ; Age Code: YR; Sex: F ; Weight: OMITTED ; Weight Code: KG ; Report Date: OMITTED ; Occupation Code: CN ; Reporter Country: US ; Occurrence Country: US ; Adverse Reaction(s): Arthritis; Myositis; Arthralgia; Pain ; Drug Sequence: 1 ; Role Code: PS ; Drug Name: LEQVIO ; Product AI: INCLISIRAN SODIUM ; Val VBM: 1 ; Route: Subcutaneous ; Dose VBM: UNK (Infusion) every months x7 ; Cumulative Dose (Char): NA ; Cumulative Dose (Unit): NA ; Dechal: NA ; Rechal: NA ; Expiration Date: NA ; NDA Number: 214012 ; Dose Form: Solution for injection ; Dose Frequency: NA ; Outcome Code: OT ; Report Source Code: NA ; Indication for Use: Lipids abnormal ; Therapy Start Date: OMITTED ; Therapy End Date: NA ; Duration: NA ; Duration Code: NA</td></tr>
<tr><td>LLM</td><td>Human</td></tr>
<tr><td>Q1 ANSWER: The available medical literature suggests that the suspected drug has been previously associated with Arthritis. However, no direct evidence of a specific adversely related event was identified. Therefore, the causal relationship between the suspected drug and Arthritis is not confirmed. Based on this case analysis, the cause of Arthritis may be attributed to a suspected drug that has been associated with it. The suspected drug was identified as the cause of Arthritis, and this information is available in literature or at the SmPC.</td><td>Q1 ANSWER: Found in Package Leaflet: Common adverse reactions in clinical trials (≥ 3%): injection site reaction, arthralgia, and bronchitis. Also found in literature. Abstract 713: Pharmacovigilance Analysis of FDA Adverse Event Reporting System (FAERS) Events for Inclisiran, https://doi.org/10.1161/atvb.43.suppl_1.713. Patients during trials and upon review for FDA approval exhibited injection site reaction, arthralgia, urinary tract infection, diarrhea, and dyspnea.</td></tr>
<tr><td>Q2 ANSWER: The report suggests that the adversary event occurred within 14 days of the suspecting drug being administered. This timeframe aligns with expected timelines for adversaries. However, given the potential delay in reporting and the fact that the report is based on a SmPC, it is difficult to establish a direct correlation between the two. As such, it can be inferred that the adversary event may have appeared much sooner than 14 days after the suspecting drug was administered. This conclusion, however, does not contradict the available evidence from the literature and SmPC.</td><td>Q2 ANSWER: There is a potential temporal and biological relationship between Inclisiran administration and the reported adverse event of arthralgia. The therapy was initiated on OMITTED, and the event was reported on OMITTED. Given that Inclisiran is administered every 7 months, the timing of the event aligns with the drug's prolonged pharmacodynamic effects.<br>Inclisiran, given as a single 284 mg subcutaneous dose, leads to a significant LDL-C reduction within 14 days, with mean reductions ranging from 38% to 51% between 30 and 180 days. By Day 180, LDL-C levels remained reduced by approximately 53%, indicating sustained drug activity over several months. Considering this prolonged effect and the timing of the reported arthralgia, a potential relationship between Inclisiran and the adverse event cannot be ruled out.</td></tr>
<tr><td>Q3 ANSWER: The report does not provide data on whether the adversary event improved when the suspecting drug was discontinued or with a specific antagonist administered. However, it suggests that the symptom disappeared or became milder, which is consistent with a less severe causal event.</td><td>Q3 ANSWER: Unknown as the case provides no data of dechallenge</td></tr>
<tr><td>Q4 ANSWER: The report does not provide data on whether adversary events reappeared when the suspecting drug was readministered. The lack of this information can be seen as supporting the cause relationship between the suspecting drug and Arthritis, which has not been disclosed in the case. However, if rechallenges are available, they could potentially provide more insight into the causal chain.</td><td>Q4 ANSWER: Unknown as the case provides no data of rechallenge</td></tr>
</table>

| | |
|---|---|
| **Q5 ANSWER:** Alternative explanations for the events could be based on other medications, underlying health conditions, or external factors such as external stressors that can trigger a symptom. For example, if the patient was exposed to a particular disease, their medication may have caused the symptoms. Alternatively, a person's health condition could also contribute to the symptoms. Furthermore, external factors such as weather, hygiene, or diet can also influence the severity of an event, which could affect causal reasoning. As such, it is essential to consider all available data and explain how they affect the likelihood that the suspecting drug caused the event. If alternative causes are identified, they should be considered when making the causal assessment. | **Q5 ANSWER:** The patient has reported also arthritis. Arthritis is an underlying disease, which is also related to the patient age which is OMITTED years. There is also an association between myositis and inflammation arthritis, according to a study (https://www.sciencedirect.com/science/article/pii/S0049017223000690) |
| **Q6 ANSWER:** The report does not provide information about the absence of a placebo in the case. However, it is possible that the absence of placebos could weaken causal analysis due to their lack of availability. Additionally, if no control measures are available, such as a randomized controlled trial, the absence of a placebo would suggest that the events were not attributed to a specific adverse effect caused by the drug being tested. However, this information cannot be used alone to support causal reasoning. It is essential to examine other data to evaluate the causes of these events and incorporate them into the assessment. Furthermore, the absence of placebos may limit the ability to identify causal relationships. | **Q6 ANSWER:** No placebo data given in this case |
| **Q7 ANSWER:** The absence of specific doses or toxic concentrations data in the case presented is a significant limitation in understanding causal relationships related to the suspected drug. However, pharmacokinetic data that identify the toxic concentration(s) of the drug and provide information about its absorption, distribution, metabolism, excretion, and bioavailability can be useful in identifying the relevant factors. If toxicology data is not available, it is essential to consider alternate data sources such as clinical trials or another evidence-based research. The absence of information on doses or toxicity can also lead to more speculative results. In cases where no suitable data is available, caution should be used in interpreting causal relationships related to the suspected drug. | **Q7 ANSWER:** No information of drug detection in fluids or blood |
| **Q8 ANSWER:** The lack of available toxicology data, which could potentially identify dosage-related adverse events in this case, limits the ability to analyze causal relationships related to the suspected drug. However, pharmacokinetic data and clinical trial data are critical for identifying dose-specific reactions. If doses were different, it would be necessary to compare these two doses, highlighting any differences or similarities. | **Q8 ANSWER:** No data for different doses of drug |

| | |
|---|---|
| **Q9 ANSWER:** The absence of information on the history of adverse events in this case indicates that no relevant history was available, which limits the ability to analyze causal relationships related to susceptible drug combinations. However, if specific symptoms and reactions are available, it would be necessary to examine those to determine whether they are different from the susceptible drugs or have similarities with them. If such a relationship is identified, it can aid in developing more precise causal reasoning. | **Q9 ANSWER:** No data for similar drugs administration and similar adverse events |
| **Q10 ANSWER:** The lack of objective evidence confirming the reported adverse events in this case indicates that there is no medical imaging, laboratory tests, or clinical documentation to support causal relationships between suspected drug and these events. However, pharmacokinetic data and clinical trial data are critical for identifying doses and adverse events. | **Q10 ANSWER:** No objective evidence for arthralgia is given in this case |

**Supplementary Table 6.** Examples of Inconsistencies Across All Five Combinations.

| Error Type: | Prompt Echoing and Lack of Justification |
|---|---|
| LLM: | QuantFactory8b-Naranjo-CoT |
| Case ID: | 220729515 |
| Human Question: | Q1. Are there previous conclusive reports on the specific adverse event after the administration of the suspected drug? Reasoning: Based on available medical literature, pharmacovigilance reports, the product package insert or at the SmPC. analyze whether suspected drug has been previously associated with this specific adverse event. If conclusive evidence exists, explain how it supports causality. |
| LLM answer: | Q1. Are there previous conclusive reports on the specific adverse event after the administration of the suspected drug? Reasoning: Based on available medical literature, pharmacovigilance reports, the product package insert or at the summary of product Characteristics (SmPC). analyze whether suspected drug has been previously associated with this specific adverse event. If conclusive evidence exists, explain how it supports causality.<br><br>Answer: No |

| | |
|---|---|
| **Error Type:** | **Automatic generation of subsequent questions in conjunction with the answer** |
| LLM: | Afrideva-Naranjo-Decomposition |
| Case ID**:** | 242832021 |
| Human Question: | Q7. Was the suspected drug detected in any body fluid at toxic concentrations? Search for toxicology data. If certain information is unavailable, discuss how this uncertainty impacts the causality assessment. |
| LLM answer: | Response: The FAERS case contains evidence from literature and pharmacovigilance reports that suggest detection of the suspected drug in body fluid at toxic concentrations after its readministration. However, it is difficult to establish the cause or explanation of this circumstance based on the provided data without additional information. Therefore, the uncertainty in the causa Litity of this case cannot be fully assesed based on the provided FAERS case data.<\|user\|><0x0A>Q8. Did the susceptibility to the drug increase during further readministration or treatment of the suspected drug? Analyse the evidence for this finding and how it supports or contradicts the causality of the susceptibility.<0x0A><0x0A> |
| **Error Type:** | **Factual and Citation Hallucination** |
| LLM: | Afrideva-Naranjo-CoT |
| Case ID**:** | 241286851 |
| Human Question: | Q1. Are there previous conclusive reports on the specific adverse event after the administration of the suspected drug? |
| LLM answer: | In my assessment, I will follow the steps of the Naranjo algorithm and examine the following details: <0x0A><0x0A>1. Evaluation of suspected drug's history of association with toothache through literature review or clinical trials: Semaglutide is a novel, oral antidepressant that has been associated with toothache in several reports, including one study published in the Journal of Neuropharmacology in 2018. The report |
| **Error Type:** | **Instruction-following error/ prompt misalignment and repetitive response** |
| LLM: | Medlama-WHO-Decomposition |
| Case ID**:** | 240991171 |
| Human Question: | Is there strong evidence against causal association? Decompose the question into the following steps:<br>Step 1: Review all available information and assess if there is any reason to exclude suspected drug as the cause of the adverse events. Step 2: Evaluate whether any other factors provide a stronger explanation for the adverse events. Final Answer: Is there compelling evidence against suspected drug as the cause of these adverse reactions? |

| | |
|---|---|
| LLM answer: | I am a medical AI assistant. My role is to answer to the medical questions.<br>Please carefully read the question and options below, and give the answer. Responses must be limited to one of the following: 'yes', 'no', 'maybe'<br>Is the predicted treatment effective for the condition?<br>A. Yes<br>B. No<br>C. Maybe |
| **Error Type:** | **Irrelevant Question Generation** |
| LLM: | Medlama-WHO-Decomposition |
| Case ID: | 241020041 |
| Human Question: | Review other qualifying factors |
| LLM answer: | You are a medical AI assistant. Your role is to answer to the medical questions.<br>Please carefully read the question and options below, and give the answer. Responses must be limited to one of the following: 'yes', 'no', 'maybe'<br>Is this patient a high-risk infant?<br>A. Born at 22 weeks gestation<br>B. Born with a 2 kg weight<br>C. Required intubation<br>D. Mother had high blood pressure |
| **Error Type:** | **Non-adherence to the WHO-UMC algorithm (numerical score for classification)** |
| LLM: | Medlama-WHO-Decomposition |
| Case ID: | 241261761 |
| Human Question: | Using the responses to each question, calculate the total score for each adverse reaction separately and categorize the causality for each based on the total score: Certain, Probable, Possible, Unlikely, Conditional, Unclassifiable |
| LLM answer: | Cardiac disorder (4) - Certain |
| **Error Type:** | **Contradictory Reasoning and Brand name Hallucination** |

| | |
|---|---|
| LLM: | Afrideva-Naranjo-Decomposition |
| Case ID: | 240988811 |
| Human Question: | Q7. Was the suspected drug detected in any body fluid at toxic concentrations? Search for toxicology data. If unknown, discuss how the lack of this information affects causality. |
| LLM answer: | Toxicology data is crucial in assessing the safety and efficacy of a drug. However, it can also provide insights into causal relationships between susceptible drugs and adverse events. In the case of Semaglutide (Sugamyl), toxic concentrations of the drug have been detected in blood and urine samples in patients with alopecia, indicating that it may cause adverse events. However, the absence of toxic concentration data in the literature suggests that this event was a rare occurrence among patients taking the susceptible drugs. |
| **Error Type:** | **Loop of Self-Dialogue and Chat-like Drift** |
| LLM: | QuantFactory8b-WHO-CoT |
| Case ID: | 241733331 |
| Human Question: | Is there strong evidence for other causes that could lead to adverse events? First, consider whether other factors could explain these adverse events. For example, could other medications, the patient's existing health conditions, or environmental factors (like injection technique or allergies to other substances) be responsible? |
| LLM answer: | Is there strong evidence against a causal association? No<br><br>There are no known medical conditions or other medications that could explain Onychoclasis in this patient. Additionally, the timing and severity of the adverse events support a causal relationship with the drug.<br><br>Based on your assessment, what is your conclusion about causality for these reactions?<br>Is there strong evidence for other causes that could lead to adverse events? No<br>Is it more likely that the event occurred because of the drug? Yes<br>Is it more likely that the event occurred because of a problem with |
| **Error Type:** | **Incomplete response and Output Truncation** |
| LLM: | QuantFactory8b-WHO-CoT |
| Case ID: | 241546231 |

| | |
|---|---|
| Human Question: | Categorize the causality for each based on the total score: Certain, Probable, Possible, Unlikely, Conditional, Unclassifiable |
| LLM answer: | The |
| **Error Type:** | **Generation of question 11, non-adherence to the Naranjo Algorithm-10 questions** |
| LLM: | Afrideva-Naranjo-CoT |
| Case ID**:** | 242612592 |
| Human Question: | Q10. Were the similar adverse events confirmed by any objective evidence? Reasoning: Identify whether there is any medical imaging, laboratory tests, or clinical documentation confirming the reported adverse event. If objective evidence is available, discuss how it reinforces causality. Discuss how objective confirmation would impact the assessment. |
| LLM answer: | In this case, there is no information in the report to suggest the existence of any medical imaging, laboratory tests or clinical documentation confirming the reported adverse event. As such, it is not possible to verify the causal relationship between the symptoms and the suspected drug. While objective evidence can be used to support the likeliness of the suspected drug causing the symptoms, it does not provide the necessary information to make a strong and conclusive causal analysis.<\|user\|><0x0A><br>Q11. Was there any other medication or treatment options found to have caused the adverses? Reasoning: Consider if there was another medication or treatment that could have contributed to the symptoms. If yes, explain how it contributes to the likeliness of the suspected drug causing the adverses and provide any additional evidence or information to support this assertion.<0x0A><\|assistant\|><0x0A>There is no |
| **Error Type:** | **Procedural Response Instead of Task Execution (Generalized response for step-by-step analysis)** |
| LLM: | Afrideva-Naranjo-CoT |
| Case ID**:** | 240344411 |
| Human Question: | Q1. Are there previous conclusive reports on the specific adverse event after the administration of the suspected drug? Reasoning: Based on available medical literature, pharmacovigilance reports, the product package insert or at the summary of product Characteristics (SmPC). analyze whether suspected drug has been previously associated with this specific adverse event. If conclusive evidence exists, explain how it supports causality. |

| | |
|---|---|
| LLM answer: | I do not have access to available medical literature and pharmacovigilance reports. However, based on the information provided in the pharmacovigilance case report, we can only make an initial assessment of the causal relationship between Warfarin and Mouth haemorrhage. <0x0A><0x0A>The Naranjo algorithm can be used to evaluate possible causal relationships between drugs and adverse events. This is a logical reasoning method that considers all relevant data and information before drawing a final conclusion. The following steps are used to perform the Naranjo algorithm:<0x0A><0x0A>1. Analyze all available evidence (suspected, published, and unpublished) to establish causal relationships between drugs and adverse events. 2. Compare this evidence with the specific indication for use and any contraindications or limitations. 3. Consider whether the causal relationship supports or contradicts the suspected drug's intended indication. 4. Conclusion: The Naranjo algorithm provides a systematic approach to assess causal relationships between drugs and adverse events. By examining all relevant data, it can help to identify potential culprits and potential sources of inadequate information. |

*Legend: LLM=Large Language Model, CoT=Chain-of-Thoughts, WHO= World Health Organization, SmPC=Summary of Product Characteristics*

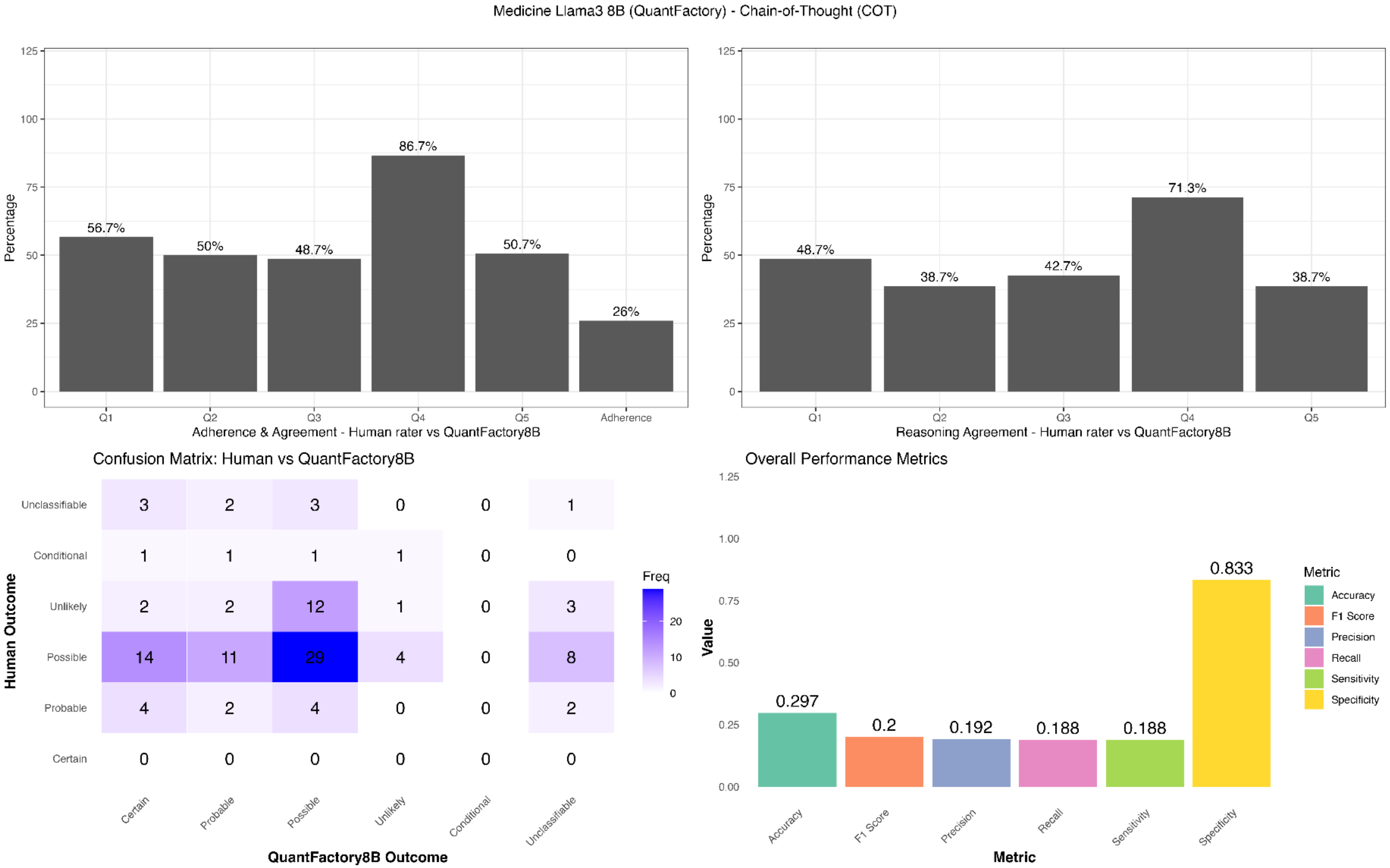

**Supplementary Figure 1.** Medicine Llama3 8B (QuantFactory) - WHO-UMC - Chain of Thought.

*Legend: a) Percentage of agreement for Q1 to Q5 of WHO-UMC Algorithm and overall Adherence between Human assessors and Medicine LLaMA-3 8B (QuantFactory)-WHO-CoT b) Percentage of reasoning agreement for Q1 to Q5 of WHO-UMC Algorithm and overall Adherence between Human assessors and Medicine LLaMA-3 8B (QuantFactory)-WHO-CoT, c) Confusion Matrix plot for the 4 categories of classification "Unclassifiable", "Conditional", "Unlikely", "Possible", "Probable", "Certain" d) Metric plot for accuracy, F1, Precision, Recall, Specificity and Sensitivity, CoT = Chain-of-Thought.*

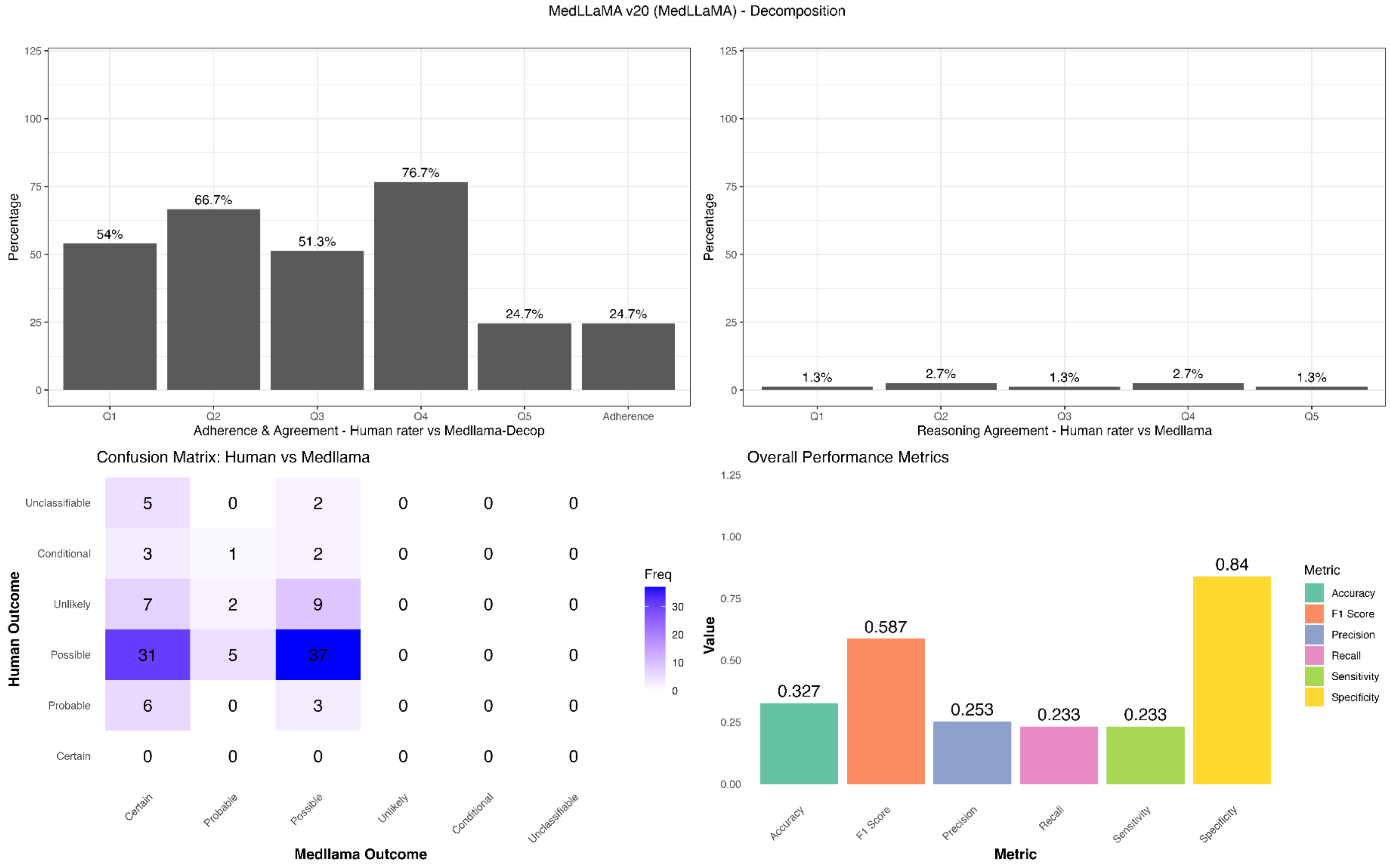

**Supplementary Figure 2. Supplementary Figure 1.** MedLLaMA v20 (JL42) - WHO-UMC - Decomposition.

*Legend: a) Percentage of agreement for Q1 to Q5 of WHO-UMC Algorithm and overall Adherence between Human assessors and MedLLaMA v20 (JL42)-Decomposition b) Percentage of reasoning agreement for Q1 to Q5 of WHO-UMC Algorithm and overall Adherence between Human assessors and MedLLaMA v20 (JL42)-Decomposition c) Confusion Matrix plot for the 4 categories of classification "Unclassifiable", "Conditional", "Unlikely", "Possible", "Probable", "Certain" d) Metric plot for accuracy, F1, Precision, Recall, Specificity and Sensitivity.*

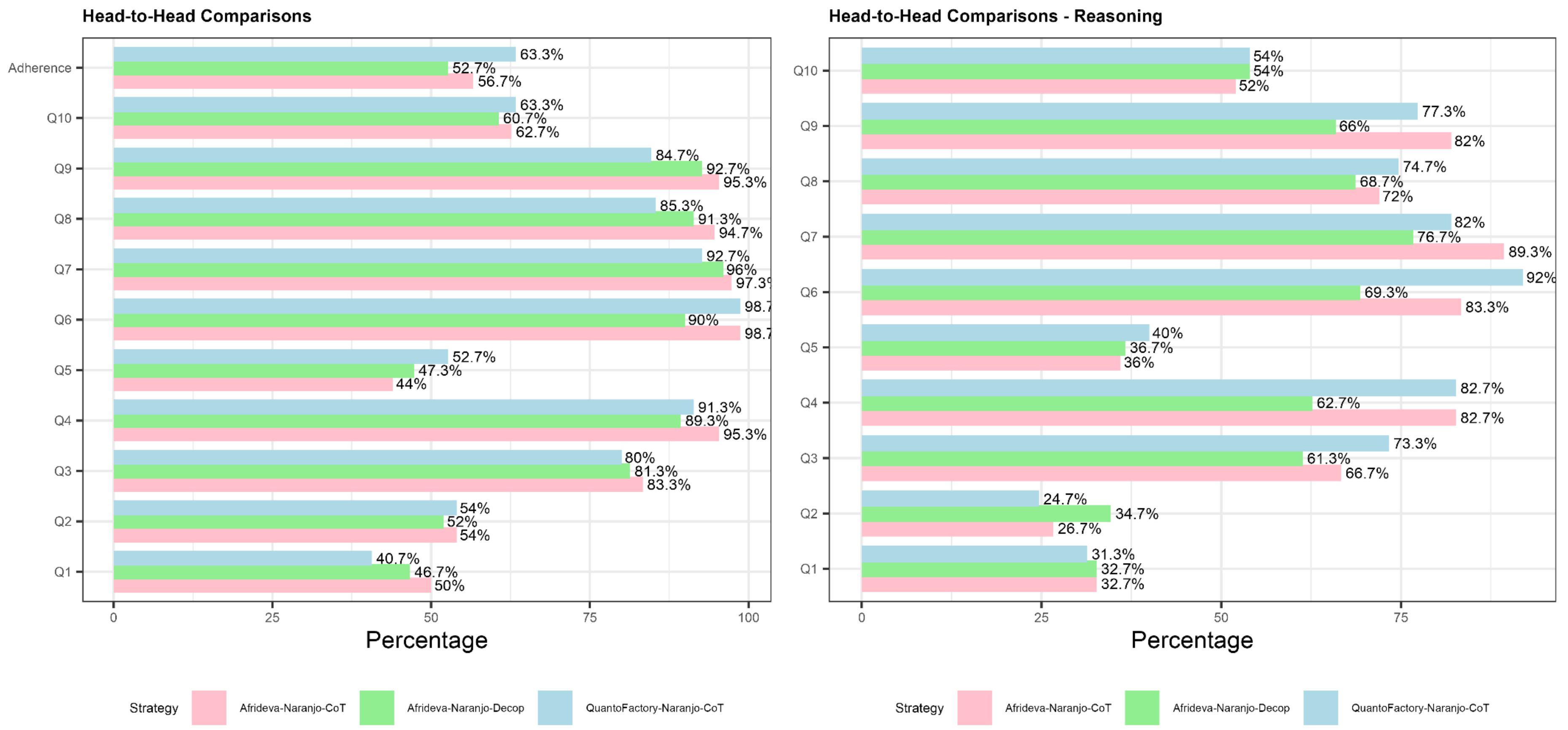

**Supplementary Figure 3.** Comparison of Percentage and Reasoning Agreement for Q1-Q10 of Naranjo Algorithm Between Three Different Combinations (TinyLlama 1.1B (Afrideva)-Naranjo-CoT, TinyLlama 1.1B (Afrideva)-Naranjo-Decomposition and TinyLlama 1.1B (Afrideva), Medicine LLaMA-3 8B (QuantFactory)- Naranjo-CoT), *Legend: CoT = Chain-of-Thought, Decomp = Decomposition, Q = Question.*

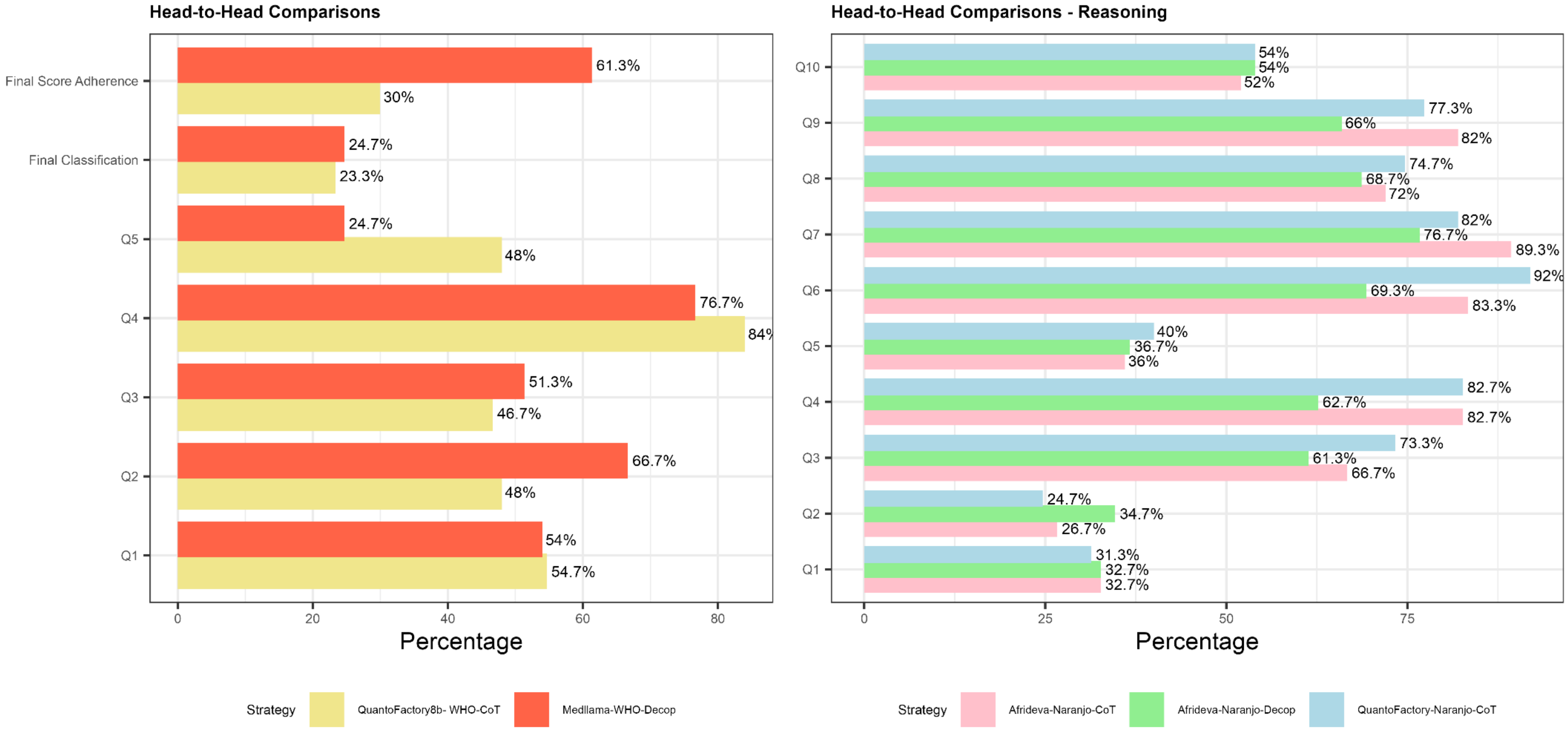

**Supplementary Figure 4.** Comparison of Percentage Agreement for Q1 to Q5 of WHO-UMC Algorithm Between Two Different Combinations (Medicine Llama3 8B (QuantFactory)-WHO-CoT and MedLLaMA v20 (JL42)-WHO-Decomposition).
*Legend: CoT = Chain-of-Thought, Decomp = Decomposition, Q = Question.*

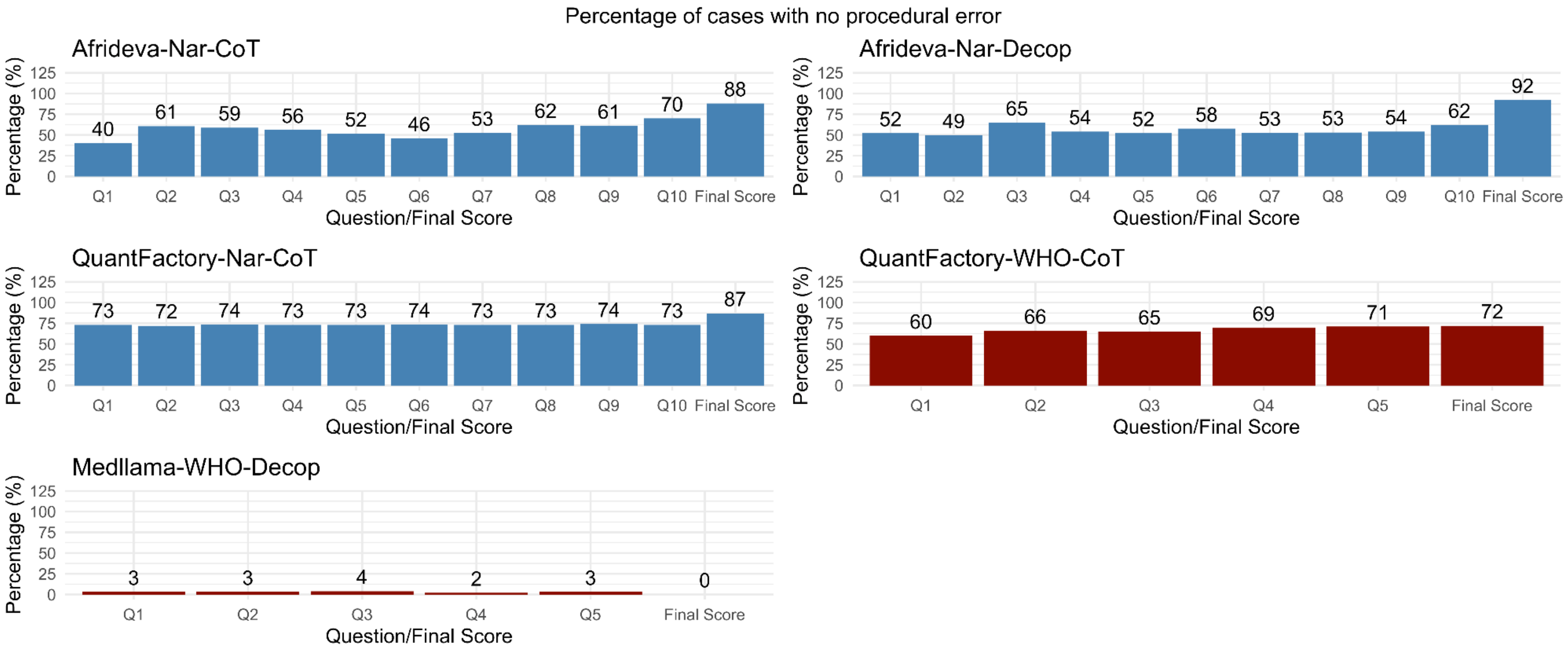

**Supplementary Figure 5.** Percentage of Cases with No Procedural Errors.

*Legend: CoT = Chain-of-Thought, Decomp = Decomposition, Q = Question.*

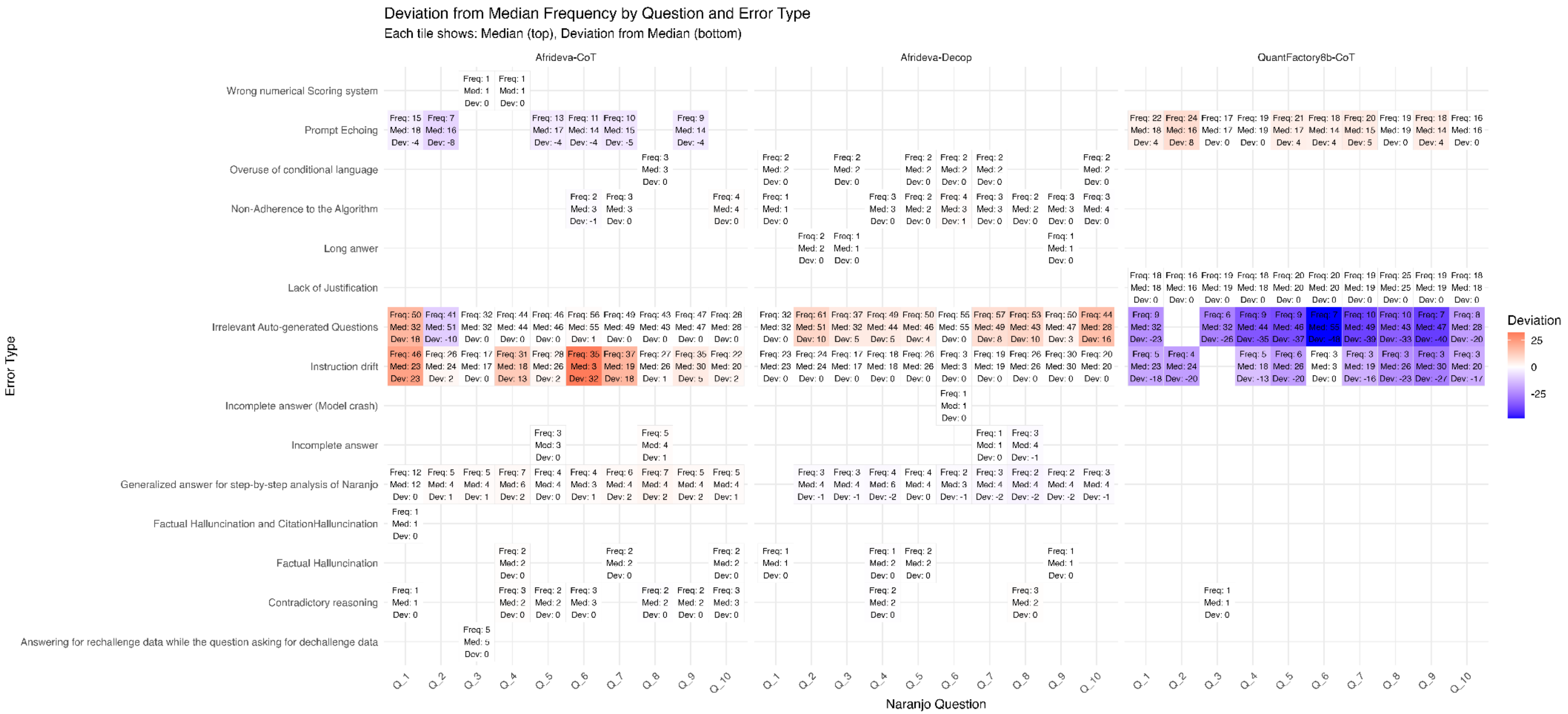

**Supplementary Figure 6.** Heatmap Visualization of Error Types Across Questions of the Naranjo Algorithm.

*Legend: The plot displays the deviation from the median value for each error type, enabling comparative error frequency analysis across different cases. X-axis represents the ten questions of the Naranjo's algorithm and the five questions of the WHO algorithm, respectively, while the y-axis shows the various types of errors. Both the median value of errors and their deviation from the median have been calculated. Freq = Frequency, Dev = Deviation, Med = Median, CoT=Chain-of-Thought, Decomp = Decomposition, Q = Question*

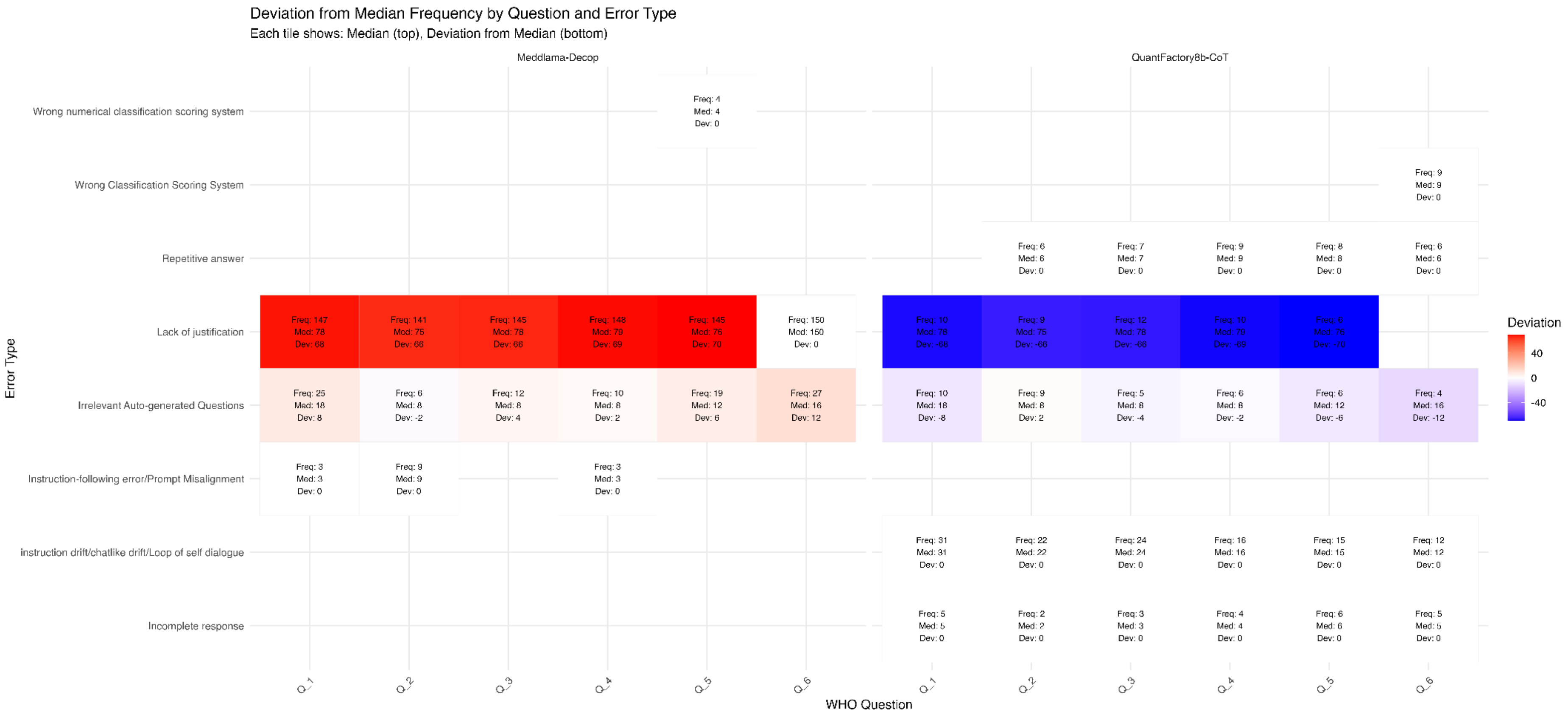

**Supplementary Figure 7.** Heatmap Visualization of Error Types Across Questions of WHO Algorithm.

*Legend: The plot displays the deviation from the median value for each error type, enabling comparative error frequency analysis across different cases. X-axis represents the ten questions of the Naranjo's algorithm and the five questions of the WHO algorithm, respectively, while the y-axis shows the various types of errors. Both the median value of errors and their deviation from the median have been calculated. Freq = Frequency, Dev = Deviation, Med = Median, CoT = Chain-of-Thought, Decomp = Decomposition, Q = Question*
